# Modeling the Effectiveness of Antibiotic Therapies Against Sepsis Using Continuous-time Hidden Markov Models

**DOI:** 10.64898/2026.07.03.26357092

**Authors:** Sophie Schmiegel, Hannah Marchi, Rainer Borgstedt, Sebastian Rehberg, Christiane Fuchs, Sina Mews

**Author notes:** both authors contributed equally (Christiane Fuchs, Sina Mews). **Corresponding author:** Christiane Fuchs Bielefeld University, Universitätsstraße 25, 33615 Bielefeld.

## Abstract

Patients suffering from sepsis need to be treated with an effective antibiotic therapy within the first hour after sepsis onset to decrease their risk of death. Microbiological data that provide information about the suitability of antibiotic therapies, however, is usually available only after 72 hours. Consequently, the treating physicians need to judge a therapy’s effectiveness based on the patients’ measured health records and their general health condition. This medical assessment is complex and requires years of experience. In our study, we investigate how statistical modeling can contribute to assessing the effectiveness of antibiotic therapies. To that purpose, we describe the effectiveness of antibiotic therapies by modeling sepsis patients’ health conditions using a three-state continuous-time hidden Markov model (ctHMM). In literature, procalcitonin (PCT) and lactate have proven to be helpful for deriving the health condition in this context. The state probabilities obtained by the ctHMM are subsequently used to quantify the effectiveness of antibiotic therapies. To this end, we apply two different approaches, namely (i) averaging of the state probabilities and (ii) a logistic regression model. For (i), we calculate the average of the state probabilities for the state indicating a sepsis-free condition over an antibiotic administration period of 48 hours. For (ii), we use the information about antibiotic susceptibility testings as dependent variable in the logistic regression model; as independent variables, we calculate the difference between state probabilities at the start of antibiotic administration and 48 hours later. With this work, we are able to better understand the relationship between laboratory values, in particular PCT and lactate, and the patients’ health condition. We further provide approaches for quantifying the effectiveness. Therefore, our work contributes to developing a clinical decision support system which helps physicians assess the effectiveness of antibiotic therapies in patients with sepsis. Supported by such a system, a physician is able to quickly adjust an ineffective therapy which avoids antibiotic resistances and increases a patient’s chance to survive a sepsis.

## 1 Introduction

Sepsis, as well as its more severe form septic shock, is a life-threatening systemic inflammatory response to an infection in the body that requires emergency treatment (Singer et al., 2016). A suitable antibiotic therapy within the first hour after diagnosis is crucial for reducing serious health consequences and lowering the risk of death (Evans et al., 2021). Pivotal data to decide on an antibiotic therapy, however, is only available around 72 hours after diagnosis depending on the hospitals’ workflows; this includes details about the sepsis-causing pathogen as well as the results of an antibiotic susceptibility testing (AST) which categorizes the suitability of antibiotic therapies into three categories: ‘susceptible (S)’ (i.e., the antibiotic therapy is suitable), ‘intermediate (I)’ (i.e., the antibiotic therapy is suitable under high dose) or ‘resistant (R)’ (i.e., the antibiotic therapy is unsuitable). At the time of diagnosis, physicians must decide on an empirical antibiotic therapy based on a paucity of data. Although clinic-specific guidelines for the empirical choice of an antibiotic therapy are available, the chosen antibiotic therapy might be unsuitable to treat the particular pathogen afflicting the patient implying that the therapy needs to be adjusted quickly. In clinical practice, the individual assessment of whether a given antibiotic therapy is effective is retrospectively made based on the course of the patient’s vital signs, laboratory values and general condition. This assessment is implicit and requires years of experience. A clinical decision support system could assist physicians in making the assessment. To develop such a decision support system, however, we need to have an explicit quantification of the effectiveness of antibiotic therapies. We define ‘effectiveness’ as the patient-individual probability that an antibiotic is suitable to treat a sepsis caused by a specific pathogen. In addition to the assessment-support, a quantified effectiveness of antibiotics is necessary in the context of health recommender systems to predict the most effective initial antibiotic for newly diagnosed patients with sepsis (Marchi et al., 2025).

Research in the field of antibiotic therapies is highly topical but particularly focuses on *antibiotic resistances* (e.g., Martínez et al., 2007; Feretzakis et al., 2021; Lewin-Epstein et al., 2021; Murray et al., 2022) and the *suitability* of antibiotic therapies for treating specific diseases (e.g., Blin et al., 2010; Logman et al., 2010; Bendala Estrada et al., 2021) as well as for treating sepsis (e.g., Burrell et al., 2016; Seymour et al., 2017; Pruinelli et al., 2018). However, to the best of our knowledge, there is little literature considering the *effectiveness* of given antibiotic therapies to patients with sepsis or septic shock. In order to assess the effectiveness of antibiotics, information on the patients’ health conditions is needed. The latter, however, is usually not available in routinely collected datasets but may be inferred from measurements on patients’ vital signs etc. In particular, continuous-time hidden Markov models (ctHMMs) are frequently used for analyzing time series data in the medical context, especially for modeling disease progression (e.g., Amoros et al., 2019; Bureau et al., 2003; Liu et al., 2015; Gultepe et al., 2014), to infer patients’ health conditions which are hidden (i.e., not directly observed) and vary over time. Specifically in the context of sepsis, Parente et al. (2018) use records of 36 patients to predict unobserved sepsis states and estimate an HMM with the two states ‘case’ (i.e., severe sepsis or septic shock) and ‘control’ (i.e., systemic inflammatory response syndrome and sepsis) for these data. The model outcome is compared to a clinical review categorization of the sepsis states. Instead of solely predicting sepsis states, however, we aim to quantify the effectiveness of administered antibiotic therapies. Rangel-Frausto et al. (1998) use a continuous-time Markov chain to model three *observed* states regarding the course of a sepsis and two absorbing states. They derive the effectiveness of an antibiotic therapy from the transition rates between the states. In contrast, our study models *unobserved* health states and derives the effectiveness of antibiotic therapies against sepsis.

To investigate the effectiveness of antibiotic therapies, we develop a two-step approach. First, we model the patients’ hidden health condition using procalcitonin (PCT) and lactate as observations to estimate a three-state ctHMM. We account for inter-individual heterogeneity by including covariates, namely age and sex, and a discrete-valued random effect (RE) in the model. Inference on the hidden states is carried out using local state decoding. Subsequently, the inferred state probabilities are used to derive the effectiveness of given antibiotic therapies. To this end, we compare two different approaches, namely (i) averaging of the state probabilities and (ii) a logistic regression model (LRM). In (i), we average the state probabilities for the healthiest state to approximate the effectiveness over the interval starting at the beginning of an antibiotic administration until 48 hours later. In approach (ii), we use the results of the AST as dependent variable in an LRM. As independent variables, we calculate the difference between the state probabilities within the time interval described above. To evaluate the applicability of the developed two-step approach using real data, we use time series data of sepsis patients of an intensive care unit (ICU) of the German hospital Evangelisches Klinikum Bethel. Our work contributes to a comprehensive understanding of the relationship between changes in a patient’s health records and the effectiveness of antibiotic therapies. This serves as a basis for building a clinical decision support system that assists physicians in assessing treatment options during the course of a sepsis and in adjusting an antibiotic therapy quickly.

This work is structured as follows: We provide medical background information underlying our modeling decisions in Section 2. In Section 3, we give an overview of the data to which we apply our two-step approach for modeling the effectiveness of antibiotic therapies. This approach is then explained in the following two sections: First, we introduce ctHMMs and our specific model for revealing the hidden health conditions of sepsis patients in Section 4; in Section 5, we then provide the methods for quantifying the effectiveness of antibiotic therapies based on the results of the previous step. Subsequently, we present and discuss our modeling results in Section 6 and conclude this work in Section 7.

## 2 Medical Background

The Third International Consensus Definitions Task Force defines sepsis as ‘life-threatening organ dysfunction caused by a dysregulated host response to infection’ (Singer et al., 2016). Consequently, it is preceded by a local infection, the course of which can worsen, ultimately resulting in sepsis (Angus and van der Poll, 2013). The most common infections are pneumonia, abdominal and urinary tract infections (Mayr et al., 2014). A septic shock is a more severe type of sepsis where, in addition to organ dysfunction, the cardiovascular system becomes severely impaired. This highly increases the probability of death (Singer et al., 2016). All in all, the mortality ranges from 30 to 50 % (Fleischmann et al., 2016). Therefore, sepsis belongs to one of the main causes of death worldwide (Singer et al., 2016).

An infection may arise from different microorganisms such as bacteria, viruses or fungi. The underlying pathogen can only be proven in the patient’s material sample. Once the pathogen has been identified, an antibiotic therapy can be initiated. Information about which antibiotic therapies are suitable is provided by an AST. However, a broad-spectrum antibiotic therapy needs to already be administered within the first hour after admission (Evans et al., 2021). As the result of an AST is usually available only after 72 hours, the decision for the first treatment and its evaluation have to be made based solely on clinical parameters and the patient’s health condition.

After therapy start, a patient’s health condition is regularly assessed to evaluate the effectiveness of the antibiotic therapy. To this end, further material samples are analyzed and the patient’s general condition is checked. In many countries, including Germany, this is carried out over 72 hours (Hagel and Brunkhorst, 2011; Roger et al., 2013). Afterwards, it is evaluated whether the patient’s health condition is noticeably better than at the start of the therapy. If so, the therapy is assessed as being effective, otherwise it is changed or escalated.

Once diagnosed, sepsis patients are closely monitored with regard to their vital signs and laboratory values. Among these are PCT and lactate, two important indicators in sepsis diagnoses, and therefore part of our study, which are explained in more detail here.

PCT is a precursor of the protein calcitonin, which belongs to the group of peptide hormones (Meisner et al., 1999). In healthy patients, a PCT of less than 0.05 ng/ml can be measured in the blood. For values between 0.05 and 2 ng/ml, a systemic infection is possible but unlikely. A PCT of 2 ng/ml up to 10 ng/ml speaks in favor of a sepsis, whereas a PCT ≥ 10 ng/ml indicates a severe sepsis or even a septic shock (Samsudin and Vasikaran, 2017). Many studies, for example Becker et al. (2008), Balci et al. (2003) and Luzzani et al. (2003), verify the importance of PCT for the diagnosis of sepsis as well as for the evaluation of the clinical course. Among others, Whang et al. (1998) pose that patients with high values of PCT tend to be sicker; they state that PCT seems to reflect a patient’s clinical course. Further, the study by Charles et al. (2009) reveals an association between appropriate empirical antibiotic therapies and a decrease in PCT.

Lactate is produced under anaerobic conditions during the metabolism of glucose (Allen and Holm, 2008). In the blood of a healthy human, a lactate level of 0.5 to 1.0 mmol/l can usually be observed (Allen and Holm, 2008). Various studies conclude that increased lactate levels are a meaningful indicator of a severe disease or injury (e.g., Bakker et al., 2013; Zhang and Xu, 2014; Aslar et al., 2004; Cerović et al., 2003; Kruse et al., 1987). Furthermore, the meaning of increased lactate levels in sepsis patients has been investigated extensively. To give a few examples, Filho et al. (2016) analyze data of a retrospective cohort study of 443 patients with sepsis or septic shock and found that a lactate level of more than 2.5 mmol/l is associated with an increased risk of death. Ryoo and Kim (2018) state that a hyperlactatemia, i.e., a lactate level of more than 2 mmol/l, is related to a poor prognosis. As a result, the Third International Consensus Definitions Task Force appoint an increased lactate level as a requirement for the diagnosis of septic shock (Singer et al., 2016). Accordingly, a hyperlactatemia speaks in favor of a severe illness and a higher mortality. Several other studies confirm these results (e.g., Trzeciak et al., 2007; Mikkelsen et al., 2009; Puskarich et al., 2012).

## 3 Data Description

In our study, we use health records of 2580 patients’ hospital stays. Each of these patients was hospitalized in the ICU of the German hospital Evangelisches Klinikum Bethel and was diagnosed with sepsis or septic shock. Based on this criterion, the patients were retrospectively selected from a period of almost twelve years (2012–2023). Since we consider all documented stays, a patient may appear twice or even more frequently in the data. However, the stays can be seen as independent events, as they are separated by at least five days without antibiotic therapy in between; we therefore use ‘stays’ and ‘patients’ synonymously in the following.

The data contains information such as patients’ core data (sex, age, height, weight, etc.), vital signs (e.g., heart rate, respiratory rate, body temperature) and laboratory values (e.g., creatinine, PCT, lactate). Data preprocessing as well as challenges arising from the data characteristics have been described in the context of a related research project (refer to Schmiegel et al. (2026)). The core data is collected once per patient and thus considered time-constant; in contrast, the vital signs and laboratory values are time-varying and measured at irregular time points. Consequently, the time points differ across the patients as well as across the different variables; thus, the available time series vary in length.

As motivated in Section 2, we consider PCT and lactate to model a patient’s health condition. We also considered other variables than PCT, but most of them led to implausible results (see Section 6.1 for more details). We thus exclude stays from the analysis for which no lactate or PCT values are available at all. Moreover, we take into account the age and sex of a patient; therefore, patients without information on age and/or sex were removed from the dataset. The age at the time of admission is considered constant over time. Since a patient’s exact time point of diagnosis is not provided, we use the start time of the first administered antibiotic therapy as suspected (approximate) time of diagnosis.

The final dataset contains four variables, namely patients’ age and sex, PCT and lactate (see Table 1 for characteristics of all variables); further, the time (in minutes) after suspected diagnosis for each measurement is provided. On average, lactate is measured approximately every five hours, whereas PCT is measured on average approximately every 2.5 days.

**Table 1:** Summary statistics for the final dataset. For age, PCT and lactate, the mean and standard deviation (sd) over all patients and time points are given, whereas for sex, the frequencies of the two categories are shown.

| Variable | Characteristic | Unit |
| --- | --- | --- |
| PCT | 9.2 (sd 20.6) | ng/ml |
| lactate | 2.4 (sd 3.5) | mmol/l |
| age | 67.5 (sd 14.8) | years |
| sex (male) | 64.6 | % |

In addition to these health records, the dataset contains reports about administered therapies for each patient, providing the antibiotic’s name as well as the time of start and end of administration. We merged overlapping administration periods and considered the treatment as combination therapy. Furthermore, the dataset provides information about 986 AST results. In our study, we use those AST results that were obtained in a time frame relevant to the therapy and in which at least one pathogen was detected. Based on the AST, we assigned effectiveness categories (‘S’, ‘I’ or ‘R’) to the given antibiotic therapies per patient. In case of combined antibiotic therapies, we assumed that they belong to class ‘S’ as long as at least one antibiotic therapy belonged to class ‘S’. In this respect, we are faced with a strong class imbalance across all patients. In particular, class ‘I’ is rarely represented. In addition, this AST result can be interpreted as ‘S’ if the dose of the antibiotic therapy is increased, but this is not considered in our study. Due to this ambiguous interpretation and the small number of stays belonging to class ‘I’, we decided to exclude this class from our data.

## 4 Continuous-time Hidden Markov Models for Modeling Patients’ Health Conditions

Our statistical approach for uncovering the unobserved effectiveness of antibiotic therapies in sepsis patients involves two consecutive steps: The first step models the patients’ health states using a ctHMM; in the second step, the resulting state probabilities are reused to quantify the effectiveness. The following sections explain the model formulation and estimation of ctHMMs, introduce our estimated baseline ctHMMs and describe the inclusion of covariates and a discrete-valued RE in the state process. The reader is referred to Zucchini et al. (2016) and Glennie et al. (2023) for further details and explanations on HMMs and to Jackson et al. (2003) for ctHMMs.

### 4.1 Primer on Continuous-time Hidden Markov Models

A ctHMM is a statistical model for time series data with possibly irregularly sampled observations at times *t* = *t*_1_, …, *t*_*T*_ and consists of two stochastic processes: An observed state-dependent process *X*_*t*_ and a hidden state process *S*_*t*_.

The state process *S*_*t*_ is modeled as an *N*-state continuous-time Markov chain. It can take on a finite number of states {1, 2, …, *N*} and fulfills the Markov property. The state process is characterized by two components, the initial state distribution 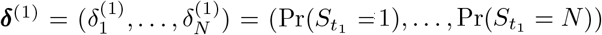 containing the probabilities of the Markov chain for starting in the respective states, and the transition rates {*q*_*gh*_}_*g≠h,g,h*=1,…,*N*_, defining the dynamics of the state process. These transition rates are summarized in the *N* × *N*-transition intensity matrix **Q**. The off-diagonal elements *q*_*gh*_ ≥ 0, *g* ≠ *h* are the rates of transitioning between the states, while the diagonal entries *q*_*gg*_ are defined as *q*_*gg*_ = − ∑ _*h*≠*g*_ *q*_*gh*_ such that the rows of **Q** sum to zero (Jackson et al., 2003). Further, the dwell time in state *g* is exponentially distributed with parameter −*q*_*gg*_. We assume **Q** to be time-homogeneous, meaning that the state dynamics remain constant over time, and can thus obtain the transition probability matrix 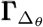 for a given time interval Δ_*θ*_ = *t*_*θ*_ − *t*_*θ−*1_, *θ* = 2, …, *T*, using the matrix exponential, namely 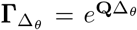 (Glennie et al., 2023). Consequently, the off-diagonal entries 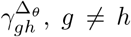, of 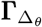 correspond to the probability of switching from state *g* to state *h* over the time interval Δ_*θ*_, accounting for an infinite number of possible state sequences in between.

Regarding the state-dependent process *X*_*t*_, the observations are allowed to be measured at irregular points in time but need to fulfill the snapshot property. This property implies that the observation of *X*_*t*_ at time *t* only depends on the state *S*_*t*_ at time *t* and not on past values of the state (Glennie et al., 2023; Patterson et al., 2017). Further, the observation at time *t* is assumed to be independent of all other observations and future or past states given the state at time *t*. Consequently, each observation follows one of *N* possible state-dependent distributions, determined by the currently active but unobserved state (Zucchini et al., 2016).

The calculation of the likelihood of all model parameters for given (univariate) observations 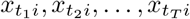 of the state-dependent process can be done using the forward algorithm (Jackson et al., 2003; Zucchini et al., 2016). In our study, *i* = 1, 2, …, *n* denotes the *i*^*th*^ patient or stay. The closed-form expression of the joint likelihood, i.e., the product of the individual likelihoods of all stays, is then given by:

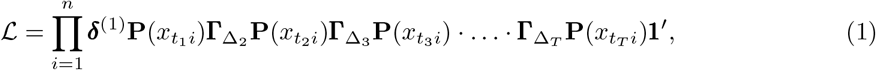

where the diagonal matrix 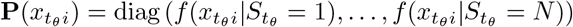 contains the state-dependent densities *f* for the observations at times *t*_*θ*_ with *θ* = 1, …, *T*, and **1**′ ∈ ℝ^*N*^ is a vector of ones. The model parameters (which are described in Section A.1 in the Appendix) are estimated by numerically maximizing the joint likelihood (1) with the help of the R package LaMa (Fischer, 2025) and automatic differentiation conveniently implemented in the R package RTMB (Kristensen, 2025).

### 4.2 Baseline Model

The considered process evolves in continuous time, with observations being measured at irregular time intervals and patients’ health conditions being hidden. Thus, we choose a ctHMM as a model for the data-generating process. Within the ctHMM, we distinguish three hidden health states: ‘non-septic state’, ‘sepsis-related state’ and ‘septic shock-related state’, in line with the Third International Consensus Definitions Task Force (Singer et al., 2016). For the state-dependent process, we use the records of PCT and lactate, i. e. 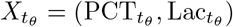 for the reasons outlined in Section 2. We assume PCT and lactate to be conditionally independent from each other given the underlying states 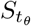, such that 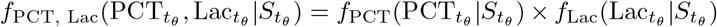. Based on the state-dependent distributions, which we assume to follow gamma distributions, we infer the underlying state process and interpret the hidden states as described above. However, the interpretation of the states is ambiguous, as it is exclusively derived from the context and the data (Patterson et al., 2017). Figure 1 visualizes our baseline model as a directed graph. The dependence structure reflects the Markov property and the observations’ conditional independence, while the unequal spacing of the observed time points illustrates the measurements’ irregularity.

**Figure 1:**
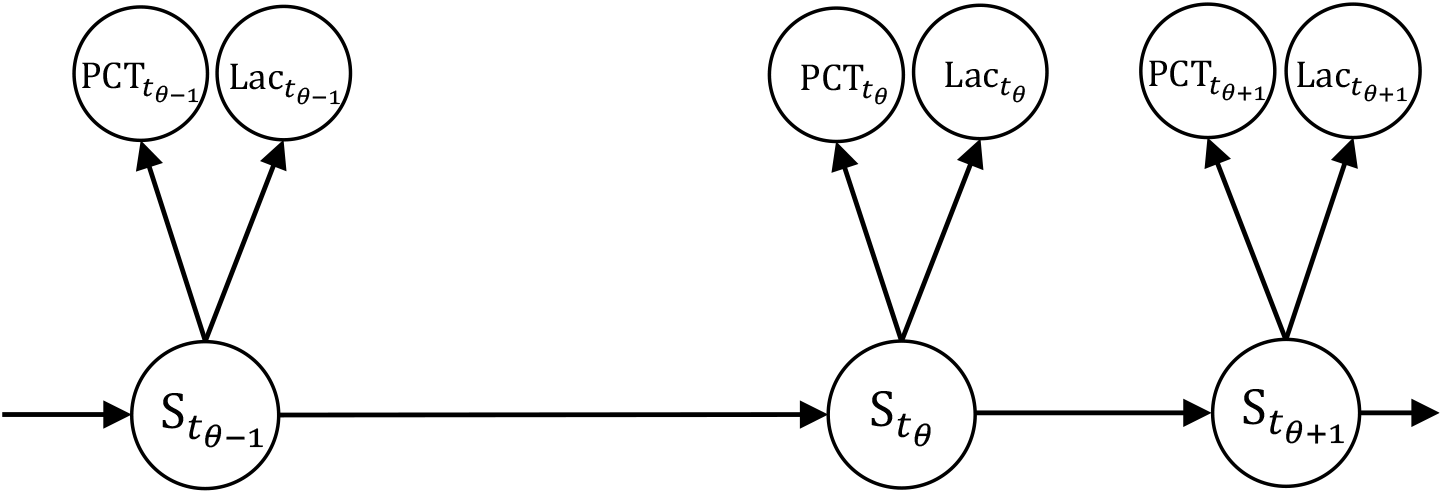
Directed graph of our baseline ctHMM with PCT and lactate constituting the state-dependent process.

### 4.3 Accounting for Inter-individual Heterogeneity

In the baseline model, we ignore heterogeneity of patients (i.e., we do not consider that patient characteristics can affect, for example, the switching behavior between the states) and therefore assume that all patients share the same parameters; consequently, we estimate one model for all stays. However, our final model allows for inter-individual heterogeneity using covariates as well as a discrete-valued RE in the state process.

We assume differences in the state-switching patterns of older and younger patients as well as male and female patients. The latter groups are usually considered separately in medical research. We extend our baseline model by including age and sex as time-constant covariates in the state process and investigate their impact on the transition dynamics. When including such subject-specific covariates in the state process, the state transition intensities are affected; we therefore model them as a function of covariates using a general linear regression framework, where the covariates serve as independent variables (Zucchini et al., 2016). Consequently, the state transition intensities for the *i*^*th*^ stay in the dataset are modeled as:

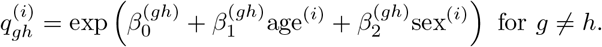

Hence, the transition intensity matrix **Q**^(*i*)^ differs across stays, depending on the patients’ age and sex, but remains constant over time.

To further account for inter-individual heterogeneity, we consider a discrete-valued RE in our model as proposed by Maruotti and Rydén (2009), McKellar et al. (2015) and Towner et al. (2016). Compared to the more commonly used continuous-valued REs, discrete-valued REs are computationally less expensive; moreover, they have the advantage that they can be interpreted as unobserved covariates (e.g., a patient’s fitness level) which separate the patients into different groups (DeRuiter et al., 2017). Following this idea, we include a discrete-valued RE in the state process and select its number of groups *K* based on the Akaike information criterion (AIC) and the Bayesian information criterion (BIC).

By considering a discrete-valued RE in the state process, we can distinguish *K* groups of patients that differ in their state-switching dynamics and thus their health condition over time. Hence, a patient’s specific transition intensity matrix **Q**^(*i*)^ is affected by an independent and identically distributed discrete-valued RE *η*^(*i*)^:

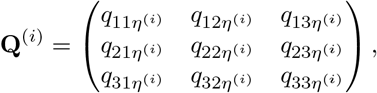

with Pr(*η*^(*i*)^ = *τ*) = *π*_*τ*_ for *τ* = 1, …, *K* and 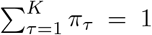, where the weight *π*_*τ*_ equals the probability that the state process of a given stay is governed by 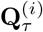 (DeRuiter et al., 2017). Taking the covariates age and sex into account, we define the elements of **Q**^(*i*)^ to be:

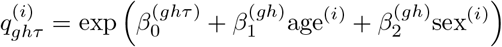

for *g* ≠ *h* and *τ* = 1, …, *K*. Figure 2 shows a graphical representation of our final model.

**Figure 2:**
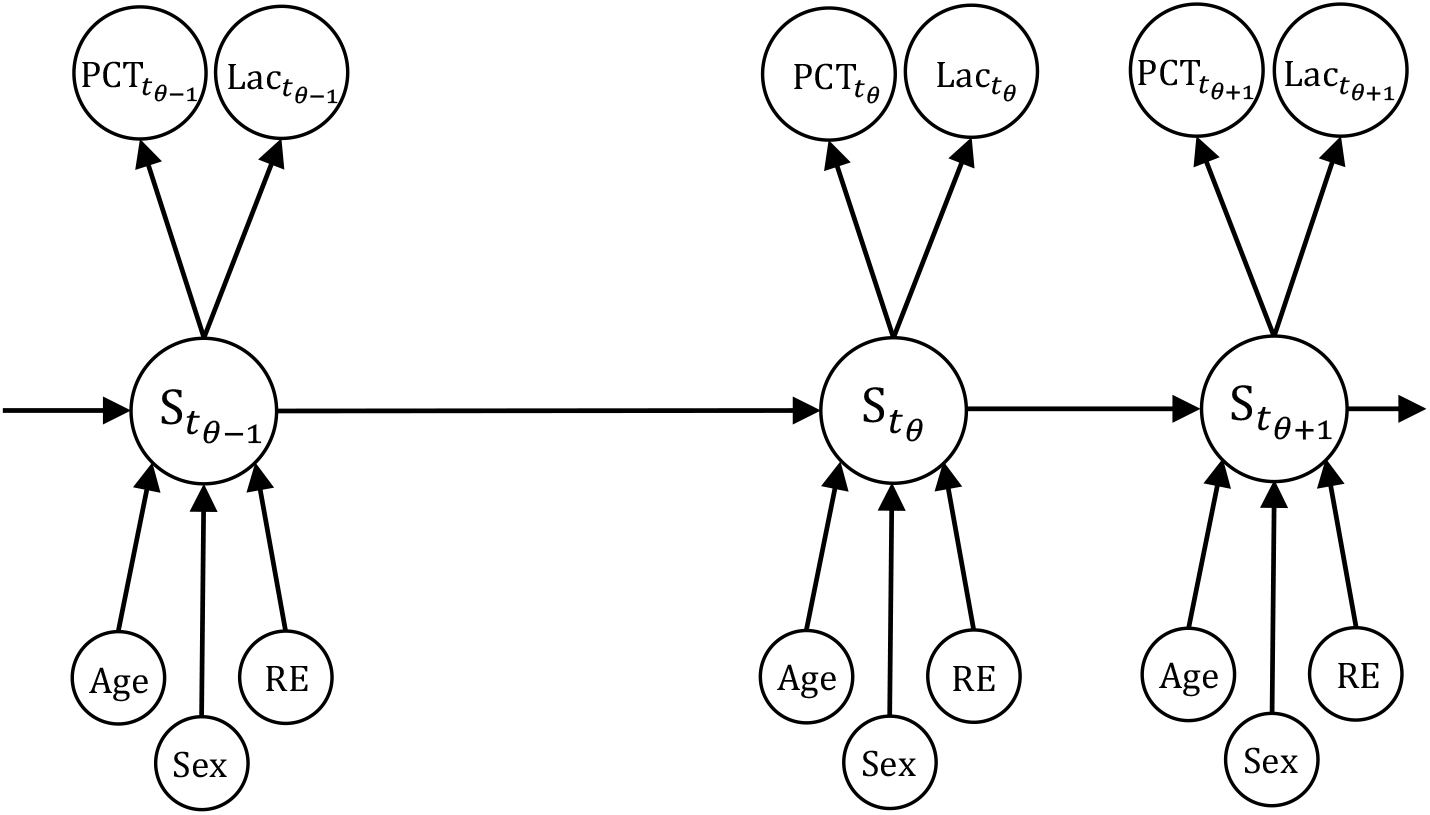
Directed graph of our final ctHMM with PCT and lactate used as observations in the state-dependent process. Age, sex and the discrete-valued RE are assumed to affect the transition intensity matrix **Q**^(*i*)^ of patient *i*.

### 4.4 State Decoding

State decoding describes the task of assigning a state to every observation of a patient’s time series. The Viterbi algorithm (Viterbi, 1967; Forney, 1973) is a common approach for decoding the states; it finds the most probable sequence of hidden states by considering the whole time series. A slightly different approach is the forward-backward algorithm (Zucchini et al., 2016), which considers each time point separately. Since the probabilities 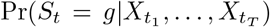 are calculated at each time point, the state *g* with the highest probability at time *t* is selected. In contrast to the Viterbi algorithm, this approach allows us to incorporate state uncertainty into the calculation of the effectiveness of an antibiotic therapy. We therefore use the forward-backward algorithm for state decoding, resulting in local state probabilities (see Zucchini et al. (2016) for details on the implementation of this approach).

## 5 Deriving the Effectiveness of Antibiotic Therapies

Like many drugs, antibiotic therapies require a certain time period until the desired effect can be achieved. In fact, they usually start to work immediately, but a measurable improvement of a patient’s health (e.g., with respect to PCT and lactate) can only be observed after several hours. Likewise, when discontinuing the antibiotic therapy, its effect extends accordingly. Figure 3 graphically visualizes this time shift. In this graphic, the start of the antibiotic therapy administration is indicated by the left-hand blue dashed line. We assume the improvement under this treatment to be delayed by about six to 24 hours; this is demonstrated by the horizontal black dashed line. Further, in clinical practice, an antibiotic therapy is reassessed after a maximum of 72 hours (gray dashed line; Hagel and Brunkhorst, 2011), taking available results of the AST into account. To aim at an earlier assessment of an administered antibiotic therapy, we consider the patients 48 hours after starting the treatment (right-hand blue dashed line) in our model and define this time interval of 0 to 48 hours as *A*_*φi*_ for every antibiotic therapy *φ* = 1, …, Φ of stay *i*. The choice of this interval is based on the idea of taking into account both measurements that are not influenced by antibiotic therapy and those that reflect the sepsis course of a patient whilst receiving antibiotic treatment. The time points of the first and the last measurements within the interval are denoted as *a*_1_ and 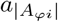, respectively, where |*A*_*φi*_| is the number of measurements in the interval *A*_*φi*_.

**Figure 3:**
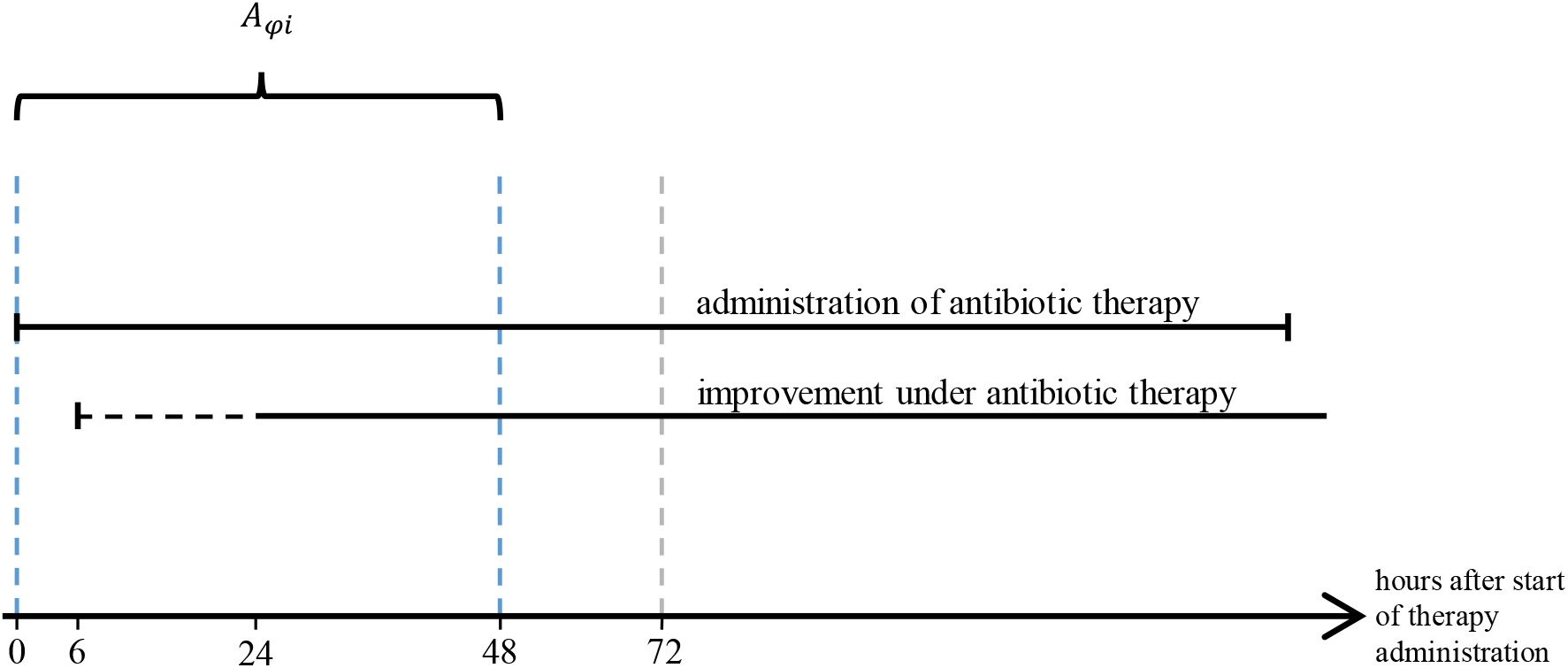
Schematic representation of the considered time interval, indicated by the blue dashed lines, for quantifying the effectiveness of antibiotic therapies. The gray dashed line marks the usual time of reassessment in clinical practice. We assume a delay in noticeable improvement under the antibiotic therapy by six to 24 hours, which is indicated by the horizontal black dashed line.

To quantify the effectiveness of antibiotic therapies, we use the local state probabilities of the non-septic state obtained by the estimated ctHMM described in Section 4. These probabilities are denoted by 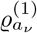, where *ν* = 1, … |*A*_*φi*_|. We consider each antibiotic therapy *φ* of each stay *I* separately and apply two different approaches, namely (i) averaging of the state probabilities and (ii) an LRM.

In approach (i), we consider 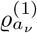 at each measurement time point *a*_*ν*_ ∈ *A*_*φi*_. The mean probability of the non-septic state for this interval, defined as

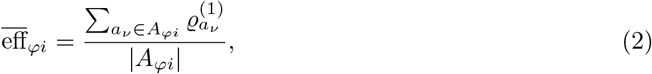

is used to approximate the effectiveness of antibiotic therapy *φ* for stay *i*. We thus consider the (mean of the) local state probabilities at each time point.

For (ii), the LRM, we compare a patient’s health condition at the start of the drug administration with the health condition 48 hours later. This is exemplarily visualized in Figure 4 using a best-case scenario for the probability curve of the non-septic state (exemplary representations of a decreasing health condition under ineffective antibiotic therapy as well as a constant health condition under an intermediate antibiotic therapy are provided in Figure A1 and Figure A2 in the Appendix). As in Figure 3, the considered time interval *A*_*φi*_ is shown using blue dashed lines. This method takes the local state probabilities of exclusively two time points into account, namely the first and the last available state probabilities 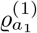 and 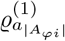, respectively. These values are visualized as yellow dots. We calculate the difference between these two probabilities (that is, the difference on the vertical axis corresponding to the change marked by the red line) and use this difference as independent variable in an LRM. We exclude the sepsis-related state and the septic shock-related state from the model due to (practical) multicollinearity. By assessing the change in the state probabilities of a patient, we emulate the exact procedure as practiced by physicians in the clinics. We do not consider further dynamics within the interval, as these can also be attributed to other events (e.g., operations) and not necessarily to the effectiveness of the given antibiotic therapy.

**Figure 4:**
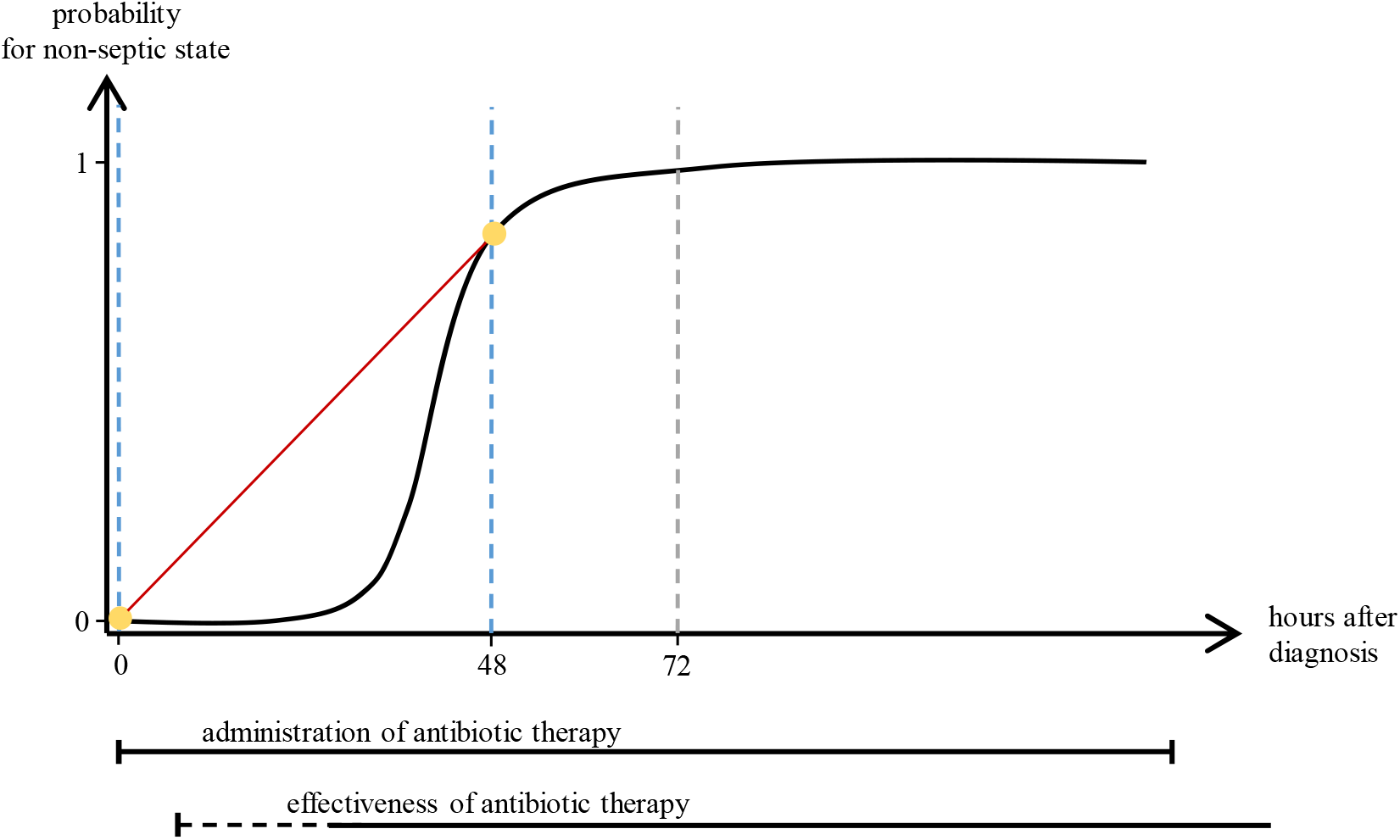
Schematic representation illustrating the idea behind the approach of using an LRM for quantifying the effectiveness of antibiotic therapies based on the difference in the local state probabilities. The black curve shows a best-case scenario for the probability of the non-septic state for a hypothetical stay over time. The two state probabilities, whose difference (in vertical direction) is included as independent variable in the model, are visualized by the yellow points.

To apply approach (ii), an antibiotic therapy can be classified as a member of one of two classes *ω* ∈ {S, R}, i.e., ‘susceptible’ and ‘resistant’. In our data, the AST provides this categorization. The classes are used as dependent variable in the LRM, which is given by

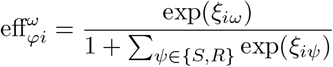

with

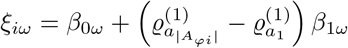

as linear predictor. For model estimation, we apply ten-fold cross-validation (CV) to account for the individuality of patients which are considered in the training data and the small sample size. We face the problem of highly imbalanced classes (see Section 3). Consequently, we handle this problem in the training data by using a combination of over- and undersampling (Shamsudin et al., 2020). We oversample the minority class and undersample the majority class, aiming at the average number of samples in a class, so that each class is equally represented in the final training set. Handling class imbalance in this way is a compromise of losing and doubling information. The outcome 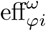 is the probability of the antibiotic therapy *φ* of stay *i* belonging to category *ω*. The resulting probability for class ‘S’ is used to approximate the effectiveness.

## 6 Results and Discussion

### 6.1 Continuous-time Hidden Markov Models

We modeled the health conditions of sepsis patients using two laboratory values, namely PCT and lactate. During model estimation, we also considered other variables than PCT (leukocytes, creatinine, interleukin 6, C-reactive protein) as observations in the state-dependent process. Most of them, however, led to implausible results, as the state-dependent distributions overlapped considerably and the estimated model parameters were inconsistent with the literature. The corresponding estimated state-dependent distributions and their parameters are provided in Section A.3 in the Appendix. Hence, based on our data, these variables seemed unsuitable for modeling the patients’ health conditions. The choice of PCT and lactate is therefore both data-driven as well as based on medical expertise; during model development, we were in close dialogue with physicians. The German Sepsis Society highly recommends to measure lactate when suspecting sepsis. Further, an increased lactate value is a necessary criterion for a septic shock. PCT is recommended for monitoring the duration of antibiotic treatment (Deutsche Sepsis-Gesellschaft et al., 2025).

We performed a series of model estimations to gradually include time-constant covariates in the state process of the ctHMM and subsequently selected the most appropriate one. To that end, we based model selection on AIC and BIC. We started by estimating the baseline model without any covariates or discrete-valued RE and include them gradually to account for individual heterogeneity. We did not consider more than four groups of the discrete-valued RE, as the computational effort increases disproportionately compared to the benefits that an additional group of the discrete-valued RE provides. Table 2 summarizes the different models as well as their AIC and BIC values. According to AIC and BIC, the model with both age and sex as covariates and four groups of the discrete-valued RE is selected (see Table 2, model 6). Hence, we choose this model as our final model and present its results in the following.

**Table 2:** Number of parameters (*p*), negative log-likelihood values (−*l*), AIC and BIC values for the six estimated models (indicated by the first column). A description of the models is provided in the second column; the superscript of discrete-valued RE indicates the number of its groups. The model selected by AIC or BIC is written in bold. ΔAIC and ΔBIC provide the difference in AIC or BIC, respectively, compared to the selected model.

| Model | Description | $p$ | $-l$ | AIC | ΔAIC | BIC | ΔBIC |
| --- | --- | --- | --- | --- | --- | --- | --- |
| 1 | baseline model | 20 | 219802.5 | 439644.9 | 3379.3 | 439851.6 | 3038.2 |
| 2 | + age | 26 | 219762.0 | 439576.0 | 3310.4 | 439844.6 | 3031.2 |
| 3 | + age + sex | 32 | 219732.9 | 439529.8 | 3264.2 | 439860.5 | 3047.1 |
| 4 | + age + sex + RE<sup>(2)</sup> | 39 | 218814.2 | 437734.5 | 1468.9 | 438828.2 | 2014.8 |
| 5 | + age + sex + RE<sup>(3)</sup> | 46 | 218445.5 | 436983.1 | 717.5 | 437458.5 | 645.1 |
| 6 | + age + sex + RE<sup>(4)</sup> | 53 | 218079.8 | **436265.6** | 0.0 | **436813.4** | 0.0 |

Figure 5 shows histograms (in dark gray) of the observed values of lactate (top) and PCT (bottom) together with the density functions of the three estimated state-dependent distributions of the chosen model in green (state one), orange (state two) and red (state three). The unconditional density is shown as black dashed lines. Each density is weighted by the proportion of time spent in the respective state derived from the local state probabilities. The estimated parameters of the state-dependent distributions and the 95 % confidence intervals (CIs) of the parameters are summarized in Table 3.

**Table 3:**
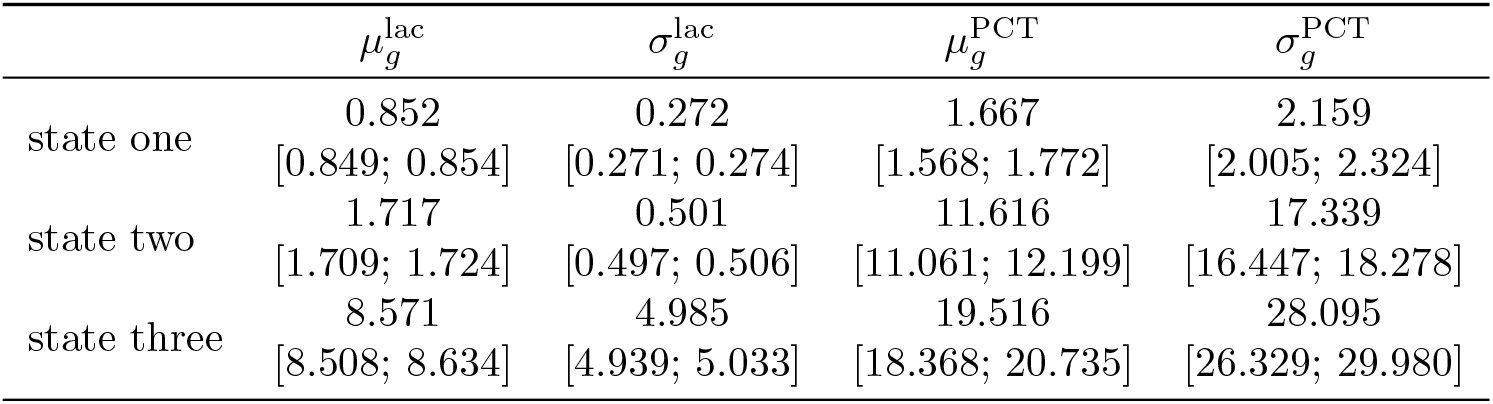
Estimated parameters of the state-dependent process. The respective 95 % CIs are given in brackets.

| | $\mu_g^{\text{lac}}$ | $\sigma_g^{\text{lac}}$ | $\mu_g^{\text{PCT}}$ | $\sigma_g^{\text{PCT}}$ |
| --- | --- | --- | --- | --- |
| state one | 0.852<br>[0.849; 0.854] | 0.272<br>[0.271; 0.274] | 1.667<br>[1.568; 1.772] | 2.159<br>[2.005; 2.324] |
| state two | 1.717<br>[1.709; 1.724] | 0.501<br>[0.497; 0.506] | 11.616<br>[11.061; 12.199] | 17.339<br>[16.447; 18.278] |
| state three | 8.571<br>[8.508; 8.634] | 4.985<br>[4.939; 5.033] | 19.516<br>[18.368; 20.735] | 28.095<br>[26.329; 29.980] |

**Figure 5:**
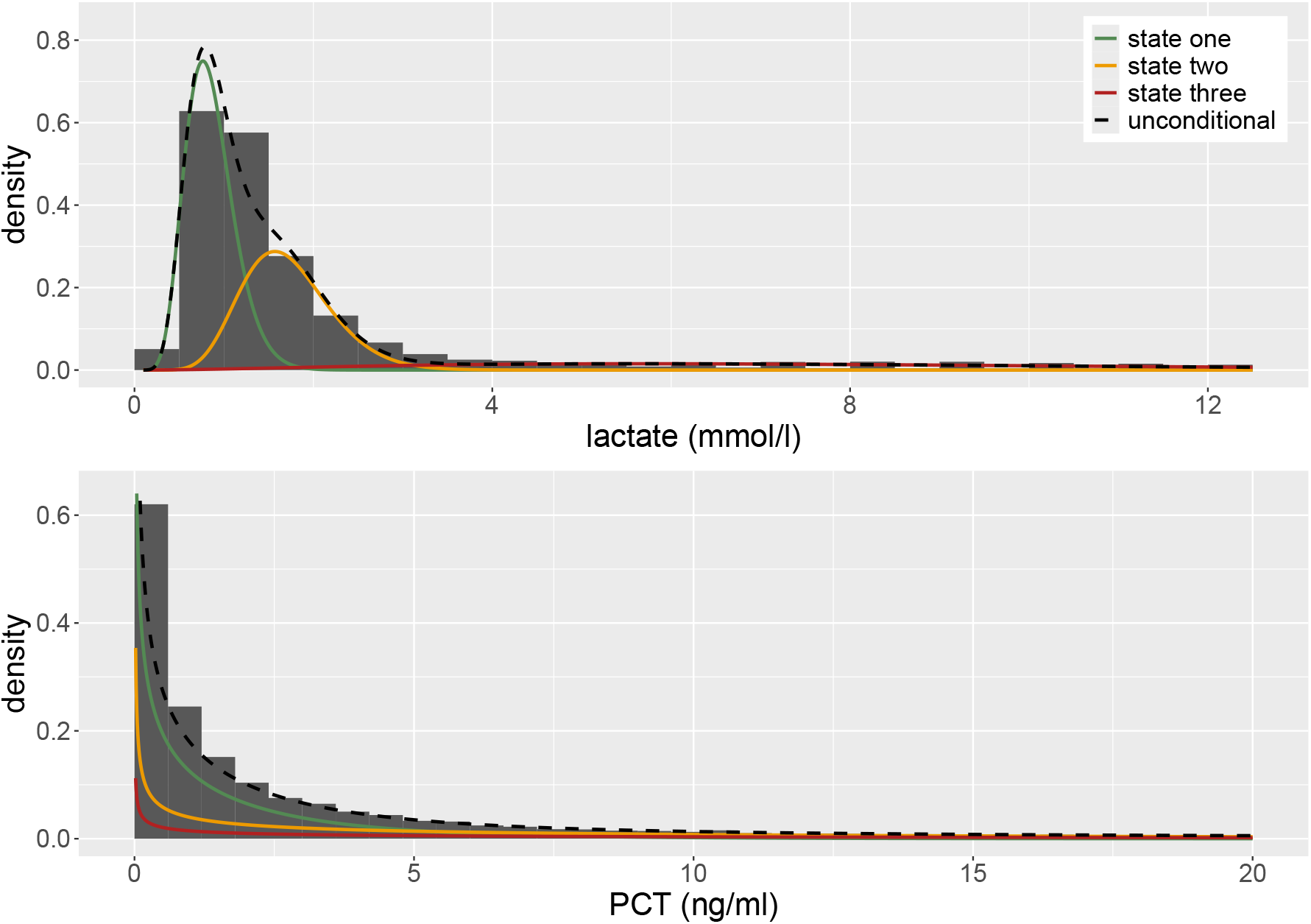
Histograms (shown in dark gray) and estimated state-dependent densities of lactate (top) and PCT (bottom) of the chosen model (see Table 2, model 6). For both laboratory values, we assume a gamma distribution. The three state-dependent distributions are weighted by the proportion of time spent in the respective state derived from the local state probabilities. The unconditional density is given as black dashed lines. Whereas typically, the gamma distribution is identified by the parameters shape *α* and scale *β*, we parametrize the gamma distribution using mean 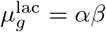 and standard deviation 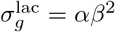 instead, due to their more intuitive interpretation.

The three states are visibly separated with respect to lactate considering the state-dependent process *X*_*t*_. In particular, state one captures small lactate values, while state two covers medium and state three mainly high lactate values. In contrast, the state-dependent distributions for PCT overlap, yet, the estimated point estimates 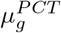 clearly differ (the corresponding CIs do not overlap) and are reasonably close to values provided in the literature (see Section 2). The overlap of the state-dependent distributions for PCT is also reflected in the estimated standard deviations 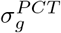 and the CIs of the parameters which are much narrower for the parameters belonging to lactate. These results are consistent with the data situation: On average, five measurements per stay are available for PCT in the data compared to 83 for lactate. Due to the small number of PCT measurements, our model is mainly driven by lactate. Lactate, however, does not react exclusively to the course of a sepsis, but also to other processes in the body. Nevertheless, our estimates convincingly align with the literature: Specifically, a systemic infection is unlikely for a PCT value below 2 ng/ml; if the PCT value exceeds 10 ng/ml, a severe sepsis or even a septic shock is suspected. These values are reflected in the means 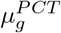 of the different states. Further, healthy patients show a lactate level of 0.5 to 1.0 mmol/l, whereas a value above 2.0 mmol/l is associated with a higher risk of death (see Section 2). Based on these results, we interpret the three states as ‘non-septic state’ (state one), ‘sepsis-related state’ (state two) and ‘septic shock-related state’ (state three), but one should keep in mind that the identification of states is completely data-driven and hence their meaning should not be over-interpreted. Particularly, it must be noted that the ctHMMs are estimated on a patient’s entire time series of PCT and lactate measured at ICU regardless of sepsis onset, as this is not included in our dataset (refer to Schmiegel et al. (2026)).

Regarding the covariates and the discrete-valued RE included in the state process, we illustrate the results based on the state transition probability matrix over six hours, **Γ**_6_, derived from the transition rates, and the dwell times in the states (i.e., the expected time spent in a state before switching to a different state). First, we turn to the discrete-valued RE. We compare **Γ**_6_ for the four discrete-valued RE groups of a mean-aged female patient:

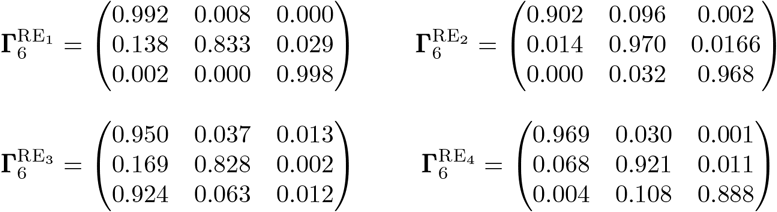

(row sums may differ from one due to rounding). The respective expected proportion of time spent in the three states for the four groups of the discrete-valued RE is provided in Table A7 in the Appendix. While patients generally tend to remain in one state over the time interval of six hours (as can be seen by the high main diagonal values), some probabilities of staying in a particular state as well as switching to a different one differ considerably across patients of the four groups. The biggest difference is visible for the probability of staying in the septic shock-related state (state three): For patients from the third group, this probability is 1.2 %, whereas it ranges from 88.8 % to 99.8 % for patients from the other groups. Consequently, patients from the third group tend to leave the septic shock-related state within six hours and most probably switch to the non-septic state; patients from the other groups, however, stay in the septic shock-related state with high probability. This behavior is also reflected in the expected dwell times summarized in Table 4. The dwell time of the septic shock-related state is 12 minutes for the first group; the other groups have a considerably higher dwell time in this state of up to 2915.7 hours. All in all, patients of the third group remain shortest in any of the states compared to the other groups, whereas patients of the first group are expected to stay particularly long in the non-septic state. In contrast, we expect patients of the first group to hardly ever leave the septic shock-related state. According to the estimated weights *π*_*τ*_, most of the patients are assigned to the fourth group; the weight estimates and the respective CIs are given in Table 5.

**Table 4:** Expected dwell times (in hours) in the three different states for the four groups of the discrete-valued RE (i.e., RE_1_ to RE_4_). The estimated regression coefficients, which are used for calculating the dwell times, are summarized in Table A5 and Table A6 in the Appendix.

|  | non-septic state | sepsis-related state | septic shock-related state |
| --- | --- | --- | --- |
| first group | 655.3 | 32.7 | 2915.7 |
| second group | 57.7 | 189.3 | 182.8 |
| third group | 16.4 | 31.2 | 0.2 |
| fourth group | 181.4 | 70.9 | 50.1 |

**Table 5:** Estimated weights for the four groups of the discrete-valued RE. Respective 95 % CIs are given in brackets.

| | $\pi_1$ | $\pi_2$ | $\pi_3$ | $\pi_4$ |
| --- | --- | --- | --- | --- |
| estimate | 0.235 | 0.252 | 0.079 | 0.435 |
| 95 % CI | [0.200; 0.273] | [0.238; 0.265] | [0.070; 0.088] | [0.419; 0.447] |

To evaluate the effect of age and sex on the state transition intensities, we compare the transition probability matrix **Γ**_6_ of the youngest and oldest male and female patients over six hours, and assume them to belong to the fourth group since most of the patients are assigned to that group (see Table 5). The transition probability matrices of these example patients result as follows (row sums may not equal one due to rounding):

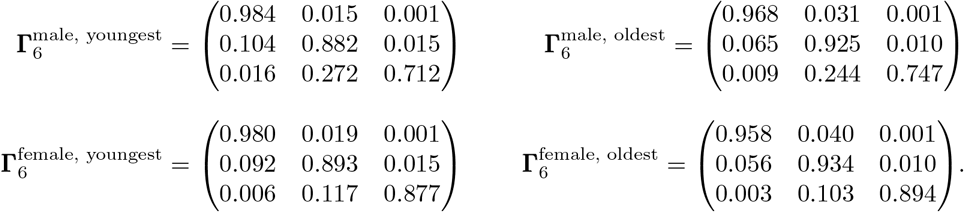

Comparing the four matrices reveals only minor differences between male and female patients regarding their switching dynamics. Such a negligible difference between the sexes is already suspected from Table 2, where the respective covariate hardly improves the model fit. A difference can be recognized in the probabilities for staying in the septic shock-related state: Female patients have a higher probability for staying in this state for six hours than male patients. Although differences are small, we keep the patient’s sex in our model as is typically done in medical contexts. Age appears to have a weak effect either; however, differences between the age groups are more pronounced when comparing the dwell times in the three states (see Table 6). While each of the four patients is expected to stay in the sepsis-related state for many hours before switching to another state, the dwell times in this state are considerably higher for older than for younger patients.

**Table 6:** Expected dwell times (in hours) in the three different states of the youngest and oldest male and female patients in our dataset assuming them to belong to the fourth group of the discrete-valued RE. The estimated regression coefficients, which are used for calculating the dwell times, are summarized in Table A5 and Table A6 in the Appendix.

|  | non-septic state | sepsis-related state | septic shock-related state |
| --- | --- | --- | --- |
| youngest male patient | 349.9 | 46.2 | 17.5 |
| oldest male patient | 175.7 | 74.4 | 20.4 |
| youngest female patient | 283.9 | 52.1 | 45.3 |
| oldest female patient | 137.4 | 85.9 | 53.3 |

Differences between younger and older as well as between male and female patients become more visible when considering the expected proportion of time spent in the three states for the four patients, which are reported in Table A8 in the Appendix. This confirms the need to take these two covariates into account.

For the other three groups of the discrete-valued RE, age and sex clearly affect the state transition intensities (the corresponding transition probability matrices and dwell times are provided in Section A.8 in the Appendix): As in case of the fourth group, younger patients stay in the non-septic state about twice as long as older patients. This holds for all three groups. In addition, younger patients belonging to the first group tend to stay in the septic shock-related state twice as long as older patients of this group. Further, for patients belonging to the first or second group, the dwell times of the septic shock-related state are roughly twice as long for female patients compared to male patients, while the dwell times of the other two states differ by a few hours. Consequently, the effect strength of age and sex depends on the discrete-valued RE group.

To decode the most likely states underlying the observations of PCT and lactate of each stay, we use local state decoding as described in Section 4.4. Figure 6 provides an example of a patient who switches several times between the three states. 24.1 % of all patients, however, do not switch between the states at all. A possible explanation is that we do not sufficiently account for patients’ individuality in our model; by assuming that all patients share the same set of parameters of the state-dependent distributions, we ignore patient-individual baseline levels of PCT and lactate. Consequently, higher values of a patient’s lactate or PCT are generally decoded as a sepsis-related state or even a septic shock-related state (see Figure A7 in the Appendix).

**Figure 6:**
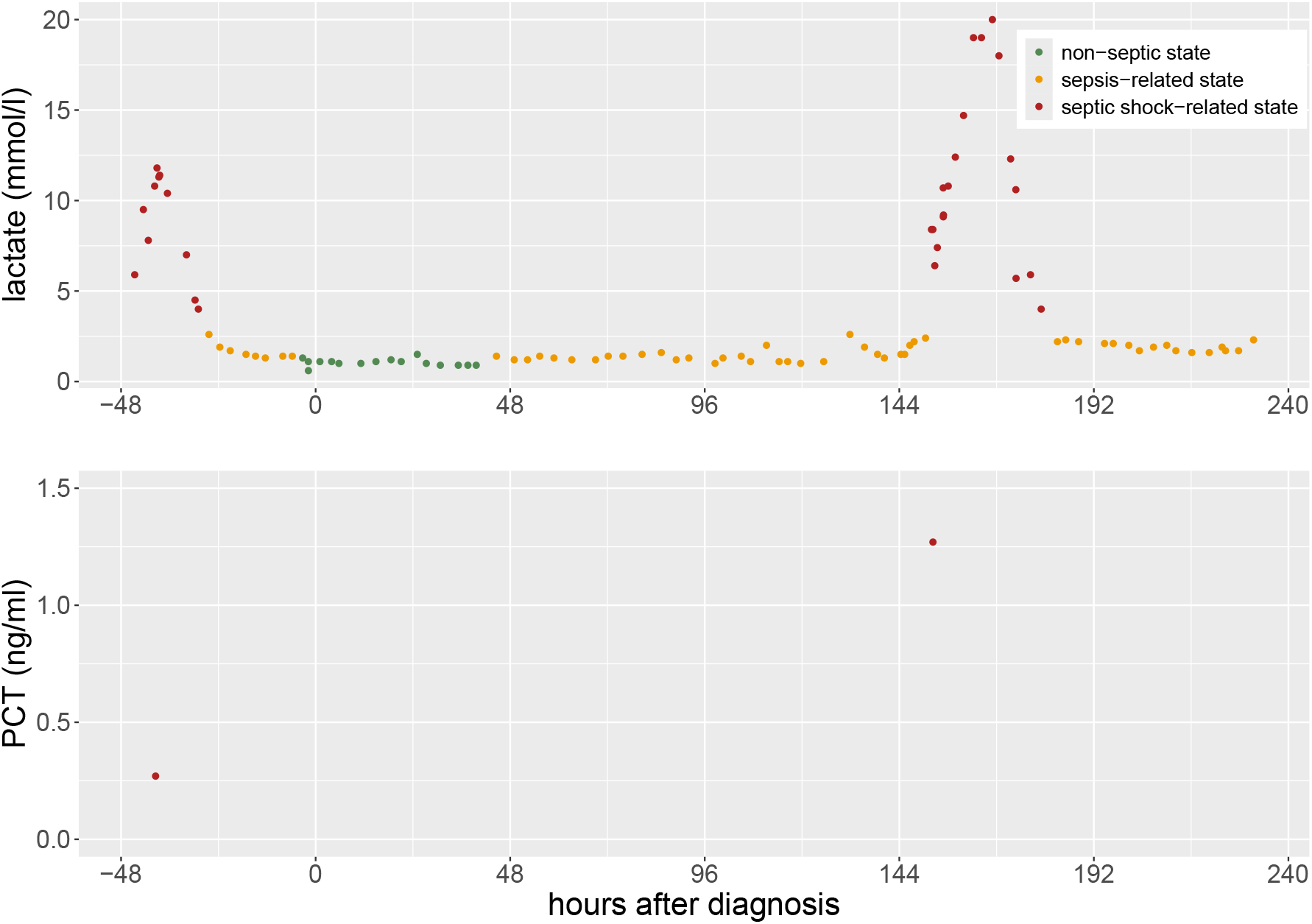
Example of a decoded time series of one stay. This patient received ‘Piperacillin/Tazobactam’ as initial antibiotic therapy and switched several times between the different states.

### 6.2 Effectiveness of Antibiotic Therapies

The ctHMM analysis estimates a patient’s course of disease, which provides indications of the effectiveness of administered antibiotic therapies via the obtained state probabilities. This estimation, however, is unsupervised. In a next step, we aim at estimating the therapies’ effectiveness in a supervised manner by combining the state probabilities with the information provided by the AST, which reflect the patient-specific ground truth about the effectiveness of antibiotics. To that end, we reuse the state probabilities obtained by local state decoding to quantify the effectiveness of 986 administered antibiotic therapies. We apply both approaches described in Section 5, i.e., (i) the averaging procedure as well as (ii) an LRM.

To evaluate the averaged state probabilities from approach (i), we relate the mean probability of the non-septic state (see Equation 2) to the results of the AST, i.e., to whether the antibiotic therapy was classified as ‘S’ or ‘R’. In a reliable real-world application, an antibiotic’s estimated effectiveness should be high for therapies of class ‘S’ and low for therapies of class ‘R’.

Figure 7 shows the antibiotic’s estimated effectiveness (horizontal axis) against the corresponding AST results (light blue for ‘S’ and dark blue for ‘R’) for all stays. The black line represents the relative frequency of ‘S’ for the respective estimated effectiveness. In the idealized case, this line would linearly increase from zero to one. The empirical line in Figure 7, however, is at a similarly high level for every estimated effectiveness. Measured across all patients, approach (i) achieves an area under the curve (AUC) of 46.7 %, an accuracy of 46.5 %, a sensitivity of 44.4 %, a specificity of 55.9 %, a precision of 81.9 % and an F1-score of 57.6 % (see Table 7). Thus, except from precision, none of these measures is acceptably high for real-world application. These results suggest that no clear conclusion can be drawn about the effectiveness of an antibiotic therapy based on the average of the state probabilities for the non-septic state.

**Table 7:** Performance of classifying a therapy’s AST result based on estimated state probabilities for the non-septic state. The latter are calculated through averaging (approach (i)) and an LRM (approach (ii)), once for the full and once for a reduced dataset. The displayed LRM performance is averaged across ten folds of CV.

| Approach | AUC | Accuracy | Sensitivity | Specificity | Precision | F1-score |
| --- | --- | --- | --- | --- | --- | --- |
| (i) | 46.7 % | 46.5 % | 44.4 % | 55.9 % | 81.9 % | 57.6 % |
| (ii), full dataset | 51.0 % | 63.1 % | 69.5 % | 34.1 % | 81.9 % | 71.7 % |
| (ii), reduced dataset | 48.0 % | 47.3 % | 43.8 % | 60.5 % | 82.2 % | 55.0 % |

**Figure 7:**
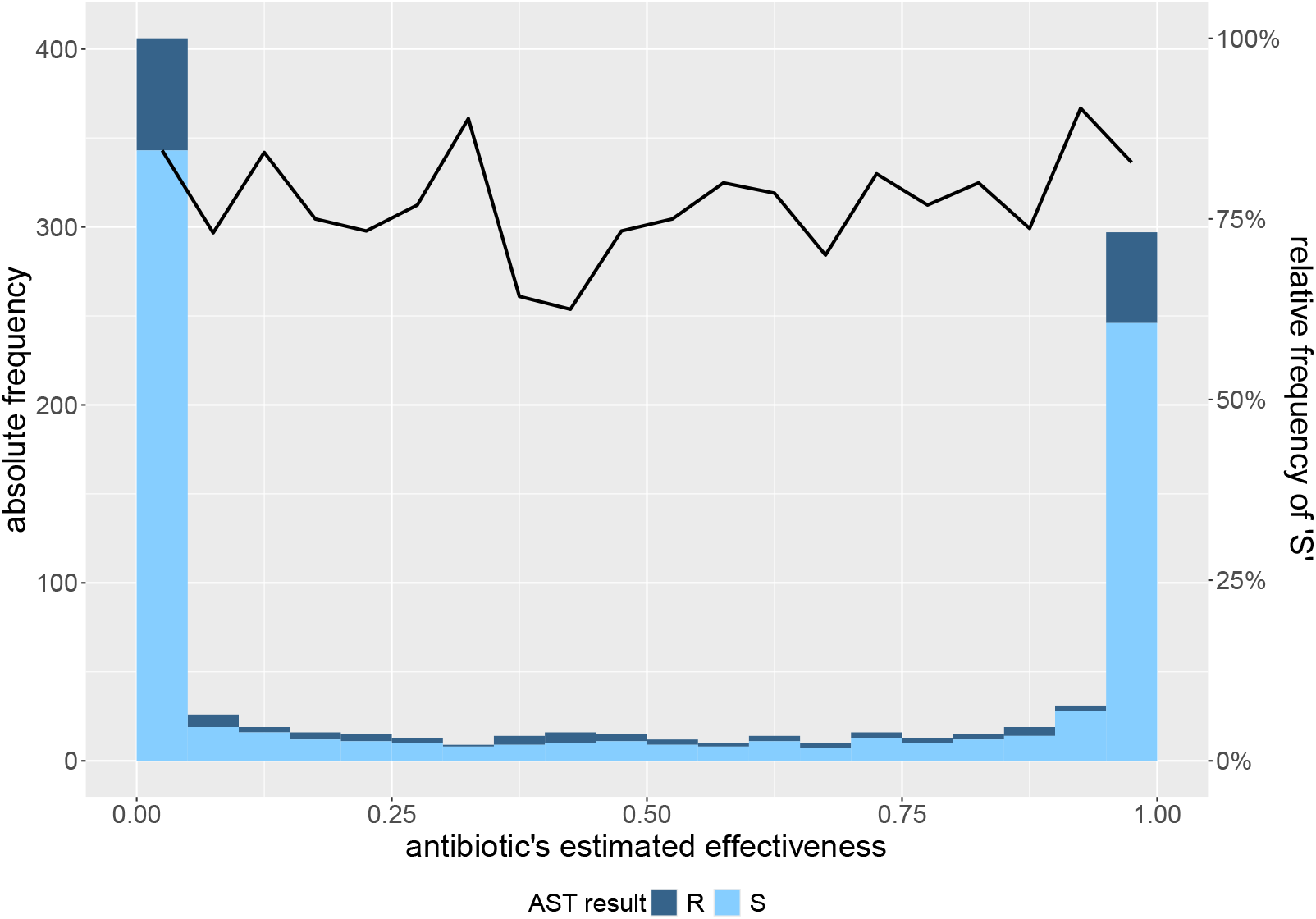
Histogram of the estimated probabilities for class ‘S’ based on approach (i) of averaging the state probabilities for the non-septic state. The colors represent the result of the AST. The black line represents the relative frequency of ‘S’ for the respective estimated effectiveness.

There are various possible reasons for this. First, this approach considers the entire time interval from 0 to 48 hours after the start of antibiotic therapy administration. Accordingly, all changes in health condition within this interval are taken into account; however, such changes are not necessarily caused by an ineffective antibiotic therapy but might be due to surgeries or other procedures which burden a patient’s body. By averaging the state probabilities of the whole interval, these unrelated events affect the calculated effectiveness. Second, it is customary in everyday clinical practice to screen a patient of poor health condition more often than a patient in good condition. Consequently, the proportion of measurements with a high probability for the non-septic state can be low, even though the given antibiotic therapy may have been effective.

The method of (ii) using an LRM to quantify the effectiveness of antibiotic therapies addresses both outlined weaknesses of approach (i): To avoid considering every single variation in health condition within the entire interval from 0 to 48 hours after the start of antibiotic therapy administration, we only consider the first and last measurements of this interval (see Section 5 for more details). This also prevents from too much weight being placed on low probabilities for the non-septic state due to frequent measurements during poor health conditions. The LRM provides probabilities for both classes ‘S’ and ‘R’. We let the probability for class ‘S’ serve as a proxy for the effectiveness of the given antibiotic therapy.

Table 7 provides empirical measures for the model’s performance on the full dataset. The model achieves high mean precision and F1-score (across ten folds of CV) of 81.9 % and 71.7 % and a medium-high sensitivity and accuracy of 69.5 % and 63.1 %, respectively. At the same time, the mean area under the curve (51.0 %) as well as the mean specificity (34.1 %) are low. These results are confirmed by the receiver operating characteristic (ROC) curve (see Figure A8 in the Appendix).

One possible explanation for the poor predictive ability of the LRM is the strong imbalance between the overrespresented class ‘S’ as compared to ‘R’ in our data, which stems from the consideration of pre-selected antibiotic combinations which come along with a higher probability of being effective. Another possible reason is that, as described above, some time series are decoded such that a patient never switches into a different state. This leads to small difference between probabilities at the start of administration and the time of reassessment. We account for the latter in the next step and investigate whether the LRM approach is nevertheless useful for patients whose health condition is estimated to change over the considered time interval.

To conduct this investigation, we reduce the data by excluding stays with 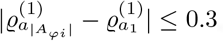, leading to a dataset of 201 stays for model estimation. Table 7 provides the resulting performance measures, showing a clear improvement in specificity with simultaneous deterioration of the other measures; only the precision remains at a high level of 82.2 %. The ROC curves show no improvement either (see Figure A9 in the Appendix). All in all, this development was to be expected because the focus on a specific group of patients came along with a substantial decrease of the sample size. In addition, the classes were still imbalanced so that class rebalancing remained necessary. Considering these limitations of the available data, the methodological approach might still provide satisfactory prediction for a larger dataset and patients whose state probabilities change over time.

## 7 Conclusion

In this study, we addressed the clinically highly relevant challenge of assessing the effectiveness of antibiotic therapy in patients with sepsis or septic shock at a time when microbiological evidence is not yet available. Because treatment decisions must be made within the first hours after sepsis onset, physicians are required to evaluate therapeutic success based solely on routinely collected clinical markers and their overall assessment of the patient’s condition. This decision-making process is complex, experience-driven, and inherently subject to uncertainty. Our objective was therefore to investigate how statistical modeling can support and formalize this early assessment of antibiotic effectiveness.

To this end, we translated the clinical evaluation process into a two-step modeling framework mimicking clinical practice: First, we represented patients’ underlying health conditions by a three-state ctHMM, using PCT and lactate as observable biomarkers and accounting for inter-individual heterogeneity through age, sex, and a discrete-valued RE in the state process. This mirrors the situation that, in clinical practice, physicians evaluate a patient’s health condition after several hours of administered antibiotic therapy compared to the initial condition. Second, we used the resulting state probabilities to quantify antibiotic effectiveness, both by averaging the probabilities of the non-septic state and by applying an LRM. This refers to the clinical practice to draw conclusions about the effectiveness of the administered antibiotic therapy. An evaluation of our modeling approach was enabled by comparing the estimated effectiveness with retrospective laboratory results, the AST.

Our analysis demonstrates that the latent effectiveness of antibiotic therapies can, in principle, be estimated using our chosen modeling approach and that this yields coherent results, in particular findings that are similar to those provided by the existing literature regarding the association with lactate and PCT (see Section 2 for a detailed literature review). While the estimated state-dependent distributions of lactate can be well separated, the according distributions for PCT are, however, less clearly distinguishable. In future studies, the ctHMM could be extended by considering patient-individual baseline levels of PCT and lactate, for example by using the clearance of the measured values. Further, and importantly, none of our approaches leads to results that largely agree with the AST in the final evaluation.

We suspect three main reasons for this: First, the considered dataset was obtained retrospectively from clinical records rather than collected specifically for the purpose our our study. It lacks important variables for modeling the course of the sepsis (e.g., the time point of sepsis onset) and for some variables such as PCT contains only a small number of observations. Such data quality issues are explained and extensively discussed in Schmiegel et al. (2026). Consequently, developing well-performing and generalizable models based on this dataset is particularly challenging. Second, patients’ health conditions do not solely depend on the effectiveness of the administered antibiotic therapy. Other factors, like surgery treatment, can additionally affect the laboratory values, especially lactate, whereas PCT is more specific to sepsis. Third, data characteristics such as non-random choices of measurement time points need to be taken into consideration. Particularly, frequent measurements at times when a patient is in a poor health condition can lead to biased results. Extending the ctHMM to account for such informative observation times, for example, by using a Markov-modulated marked Poisson process (MMMPP), might improve inference on the hidden health conditions. MMMPPs have been used to predict patients’ risk of clinical deterioration (Alaa et al., 2017) and to model disease processes based on electronic health records (Lange et al., 2015) and claims data (Mews et al., 2023).

In summary, PCT and lactate have proven to be important laboratory values, allowing us to better understand the relationship between health records and patients’ conditions using ctHMMs. To quantify the effectiveness of administered antibiotic therapies, we mimicked the procedure used in everyday clinical practice by applying statistical methods. Such methods can provide substantial gains in knowledge, provided they are based on high-quality data and that relevant data characteristics are adequately taken into account. These ideas bring us closer to a clinical decision support system which helps physicians in assessing treatment options during the course of sepsis and in promptly adjusting an antibiotic therapy. A prolonged use of ineffective antibiotic therapies could consequently be prevented, potentially reducing the risk of antibiotic resistance and increasing a patient’s chance of surviving sepsis.

## Computational Details

The whole analysis was conducted using R version 4.4.2 (R Core Team, 2024). The code is available at https://github.com/fuchslab/Effectiveness_of_Antibiotic_Therapies.

## Acknowledgments

We would like to thank Philipp Hege, Tamara Schamberger and Jan-Ole Fischer for valuable comments and stimulating discussions. Further, we thank all members of the ‘KINBIOTICS’ project.

## Funding

This study was supported by a grant from the German Federal Ministry of Health (Bundesministerium für Gesundheit), grant ZMVI1-2520DAT930 (Kinbiotics).

## Conflict of Interest

None declared.

## Ethical Considerations

Ethical approval was given by the Ethics Committee of the Westphalia-Lippe Medical Association and the University of Münster (Ethik-Kommission der Ä rztekammer Westfalen-Lippe und der Westfälischen Wilhelms-Universität Münster); case number: 2021-699-f-S.

## Data Availability Statement

Due to data privacy protection, we are not allowed to share the original data.

## A Appendix

### A.1 Model Parameters

In the context of model estimation, 53 parameters need to be estimated for the final model (i.e., the model with age and sex as covariates and four groups of the discrete-valued RE) in total.

For the laboratory values PCT and lactate, we assume a gamma distribution, which is identified by the parameters shape *α* and scale *β*. We reparameterize the gamma distribution using mean 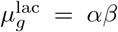 and standard deviation 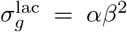 instead (similarly for PCT), due to their more intuitive interpretation. In detail, we estimate the following parameters:

- six means (one mean per state and laboratory value)
- six standard deviations (one per state and laboratory value)
- two parameters for ***δ***^(1)^
- twelve beta coefficients for the state transition intensities
- 24 coefficients for the groups of the RE
- three weights for the groups of the RE

### A.2. Exemplary Scenarios for the Course of a Patient’s Health Condition

**Figure A1:**
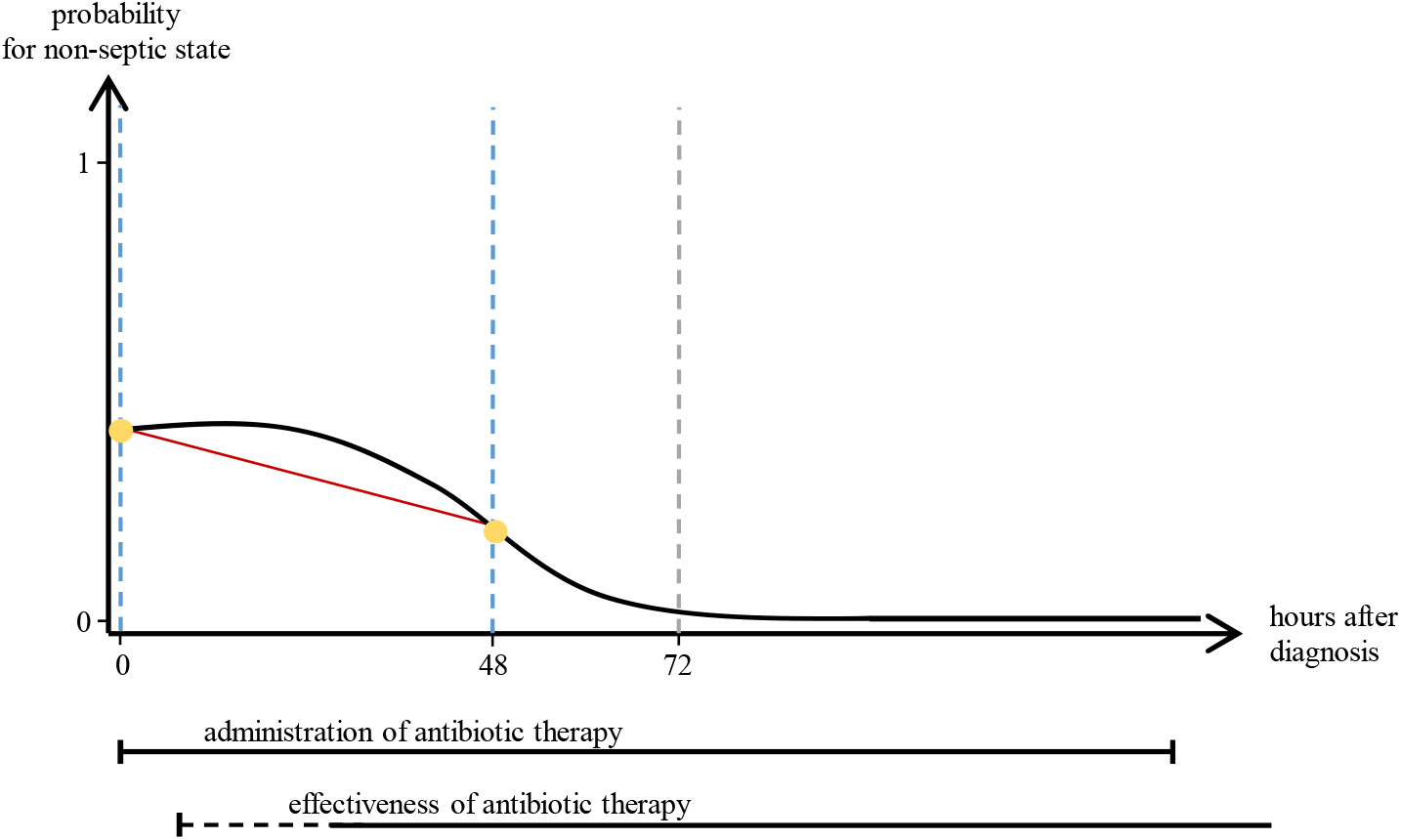
Example of a decreasing health condition under ineffective antibiotic therapy.

**Figure A2:**
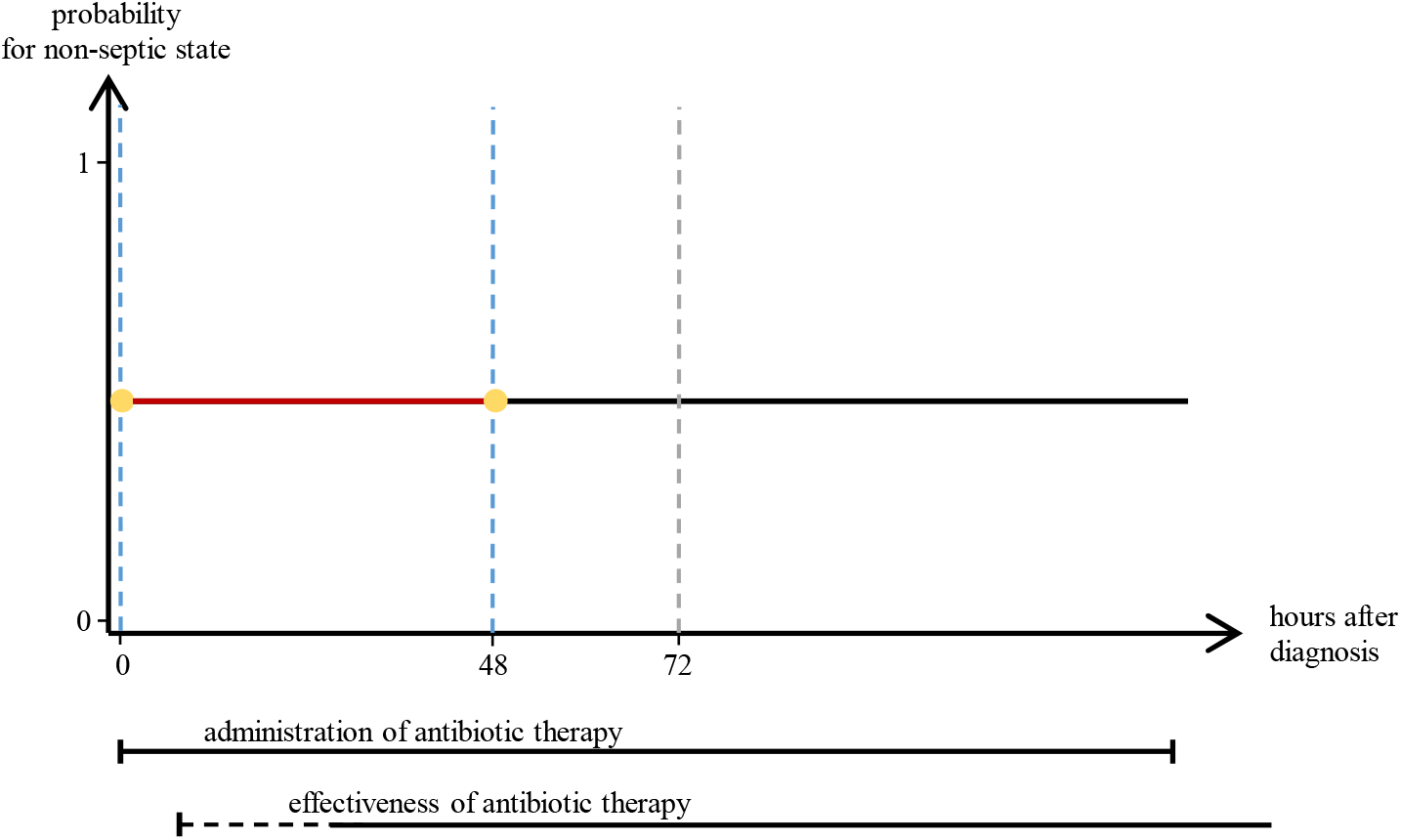
Example of a constant health condition under intermediate antibiotic therapy.

### A.3 Histograms With Estimated State-dependent Distributions

#### A.3.1 Lactate and Leukocytes

**Figure A3:**
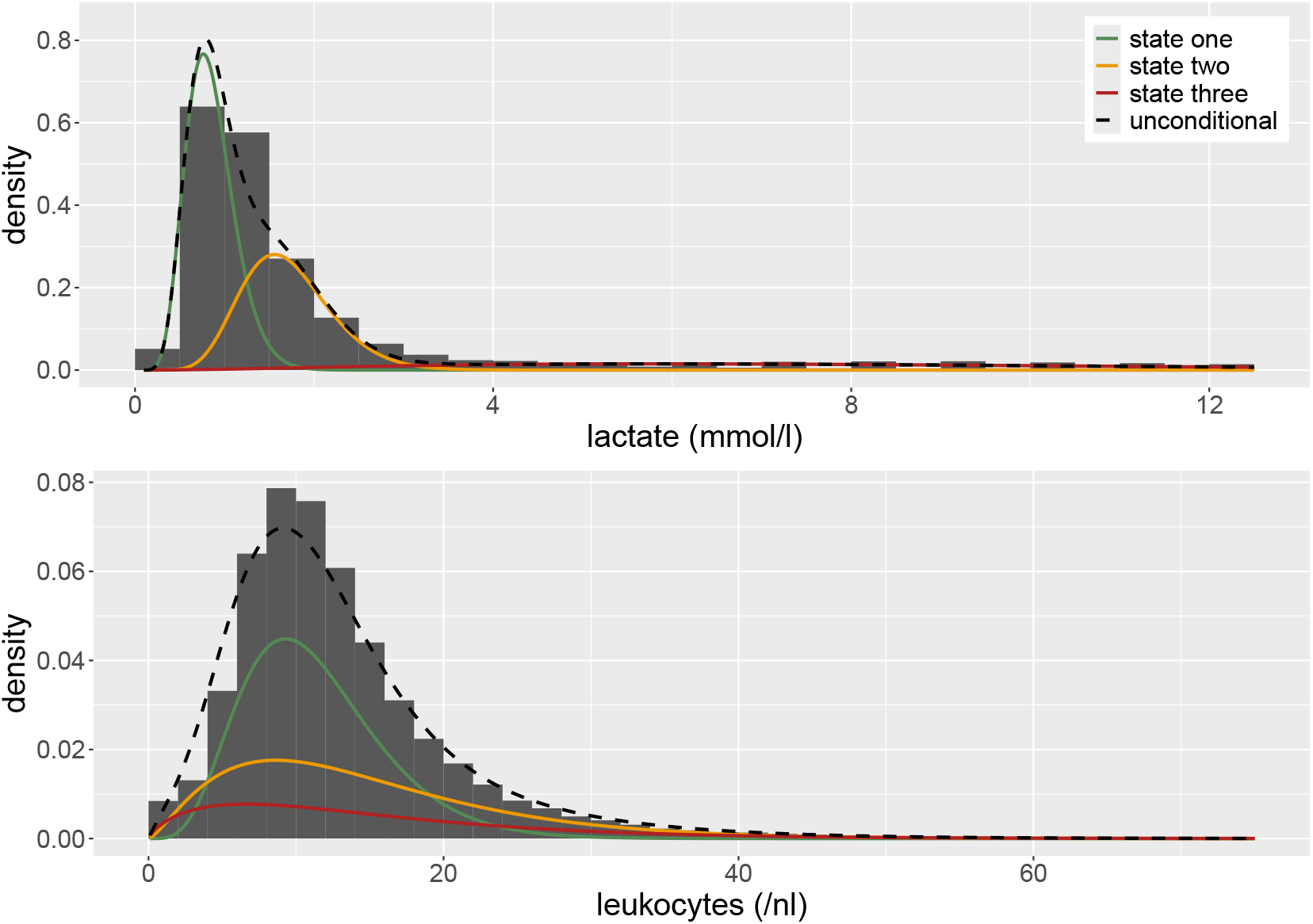
Histograms (shown in dark gray) and estimated state-dependent distributions of lactate (top) and leukocytes (bottom). For both laboratory values, we assumed a gamma distribution. The three state-dependent distributions are weighted by the proportion of time spent in the respective state derived from the local state probabilities. The unconditional density is given as black dashed line.

**Table A1:**
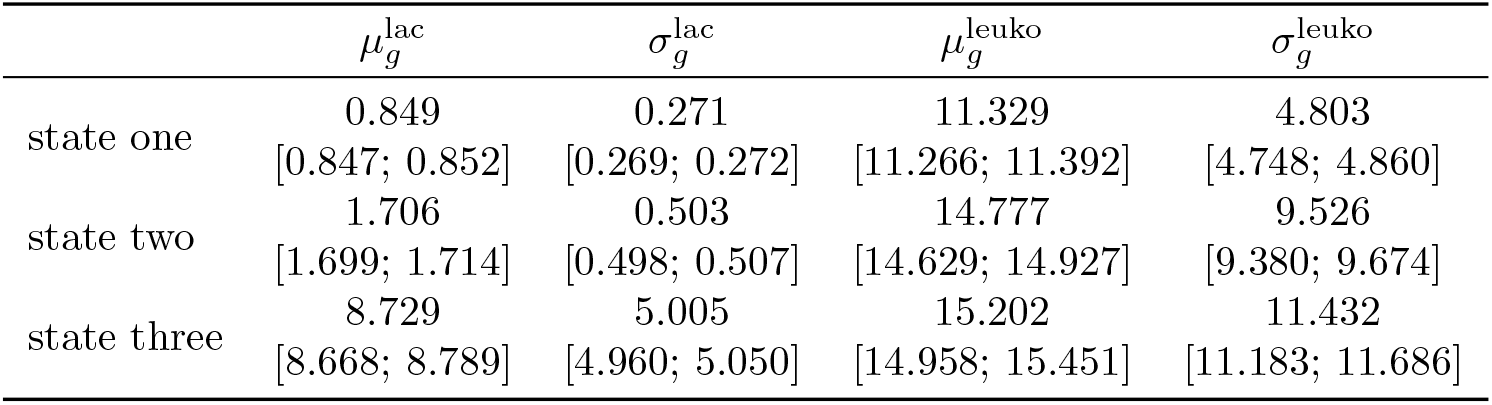
Estimated parameters of the state-dependent processes of lactate and leukocytes. Their respective 95 % CIs are given in brackets.

| | $\mu_g^{\text{lac}}$ | $\sigma_g^{\text{lac}}$ | $\mu_g^{\text{leuko}}$ | $\sigma_g^{\text{leuko}}$ |
| --- | --- | --- | --- | --- |
| state one | 0.849<br>[0.847; 0.852] | 0.271<br>[0.269; 0.272] | 11.329<br>[11.266; 11.392] | 4.803<br>[4.748; 4.860] |
| state two | 1.706<br>[1.699; 1.714] | 0.503<br>[0.498; 0.507] | 14.777<br>[14.629; 14.927] | 9.526<br>[9.380; 9.674] |
| state three | 8.729<br>[8.668; 8.789] | 5.005<br>[4.960; 5.050] | 15.202<br>[14.958; 15.451] | 11.432<br>[11.183; 11.686] |

#### A.3.2 Lactate and Creatinine

**Figure A4:**
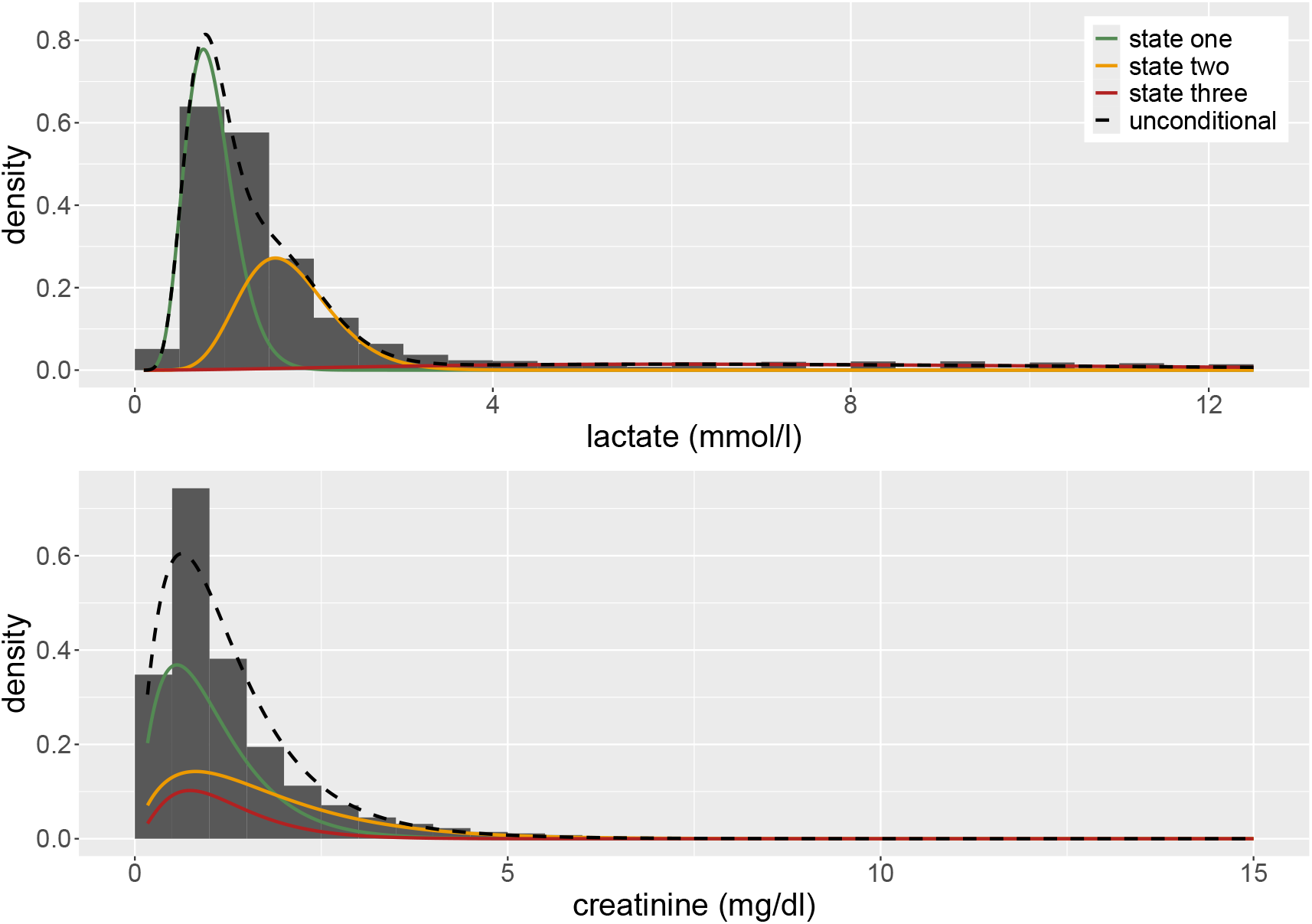
Histograms (shown in dark gray) and estimated state-dependent distributions of lactate (top) and creatinine (bottom). For both laboratory values, we assumed a gamma distribution. The three state-dependent distributions are weighted by the proportion of time spent in the respective state derived from the local state probabilities. The unconditional density is given as black dashed line.

**Table A2:** Estimated parameters of the state-dependent processes of lactate and creatinine. Their respective 95 % CIs are given in brackets.

| | $\mu_g^{\text{lac}}$ | $\sigma_g^{\text{lac}}$ | $\mu_g^{\text{crea}}$ | $\sigma_g^{\text{crea}}$ |
| --- | --- | --- | --- | --- |
| state one | 0.853<br>[0.851; 0.856] | 0.273<br>[0.271; 0.275] | 1.034<br>[1.025; 1.044] | 0.698<br>[0.688; 0.708] |
| state two | 1.722<br>[1.715; 1.729] | 0.511<br>[0.507; 0.516] | 1.719<br>[1.700; 1.739] | 1.246<br>[1.228; 1.266] |
| state three | 8.811<br>[8.751; 8.871] | 4.995<br>[4.950; 5.040] | 1.187<br>[1.168; 1.206] | 0.729<br>[0.711; 0.748] |

#### A.3.3 Lactate and Interleukin 6

**Figure A5:**
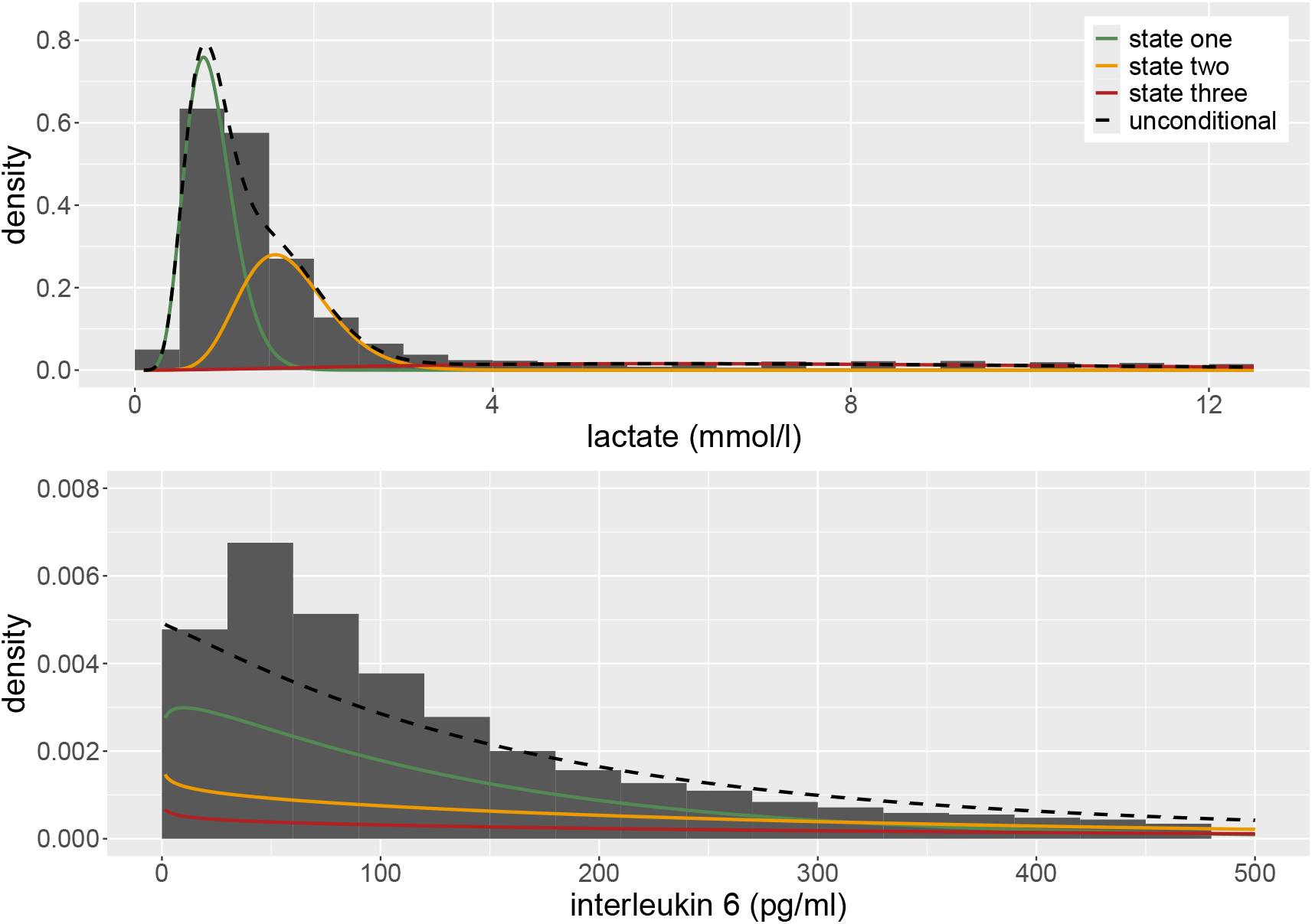
Histograms (shown in dark gray) and estimated state-dependent distributions of lactate (top) and interleukin 6 (bottom). For both laboratory values, we assumed a gamma distribution. The three state-dependent distributions are weighted by the proportion of time spent in the respective state derived from the local state probabilities. The unconditional density is given as black dashed line.

**Table A3:** Estimated parameters of the state-dependent processes of lactate and creatinine. Their respective 95 % CIs are given in brackets.

| | $\mu_g^{\text{lac}}$ | $\sigma_g^{\text{lac}}$ | $\mu_g^{\text{IL-6}}$ | $\sigma_g^{\text{IL-6}}$ |
| --- | --- | --- | --- | --- |
| state one | 0.853<br>[0.850; 0.855] | 0.270<br>[0.268; 0.272] | 140.161<br>[136.902; 143.497] | 135.051<br>[131.091; 139.130] |
| state two | 1.715<br>[1.708; 1.723] | 0.501<br>[0.496; 0.505] | 328.377<br>[320.136; 336.829] | 344.993<br>[335.155; 355.120] |
| state three | 8.766<br>[8.706; 8.827] | 5.011<br>[4.966; 5.056] | 430.087<br>[416.721; 443.882] | 462.058<br>[445.700; 479.016] |

#### A.3.4 Lactate and C-reactive Protein

**Figure A6:**
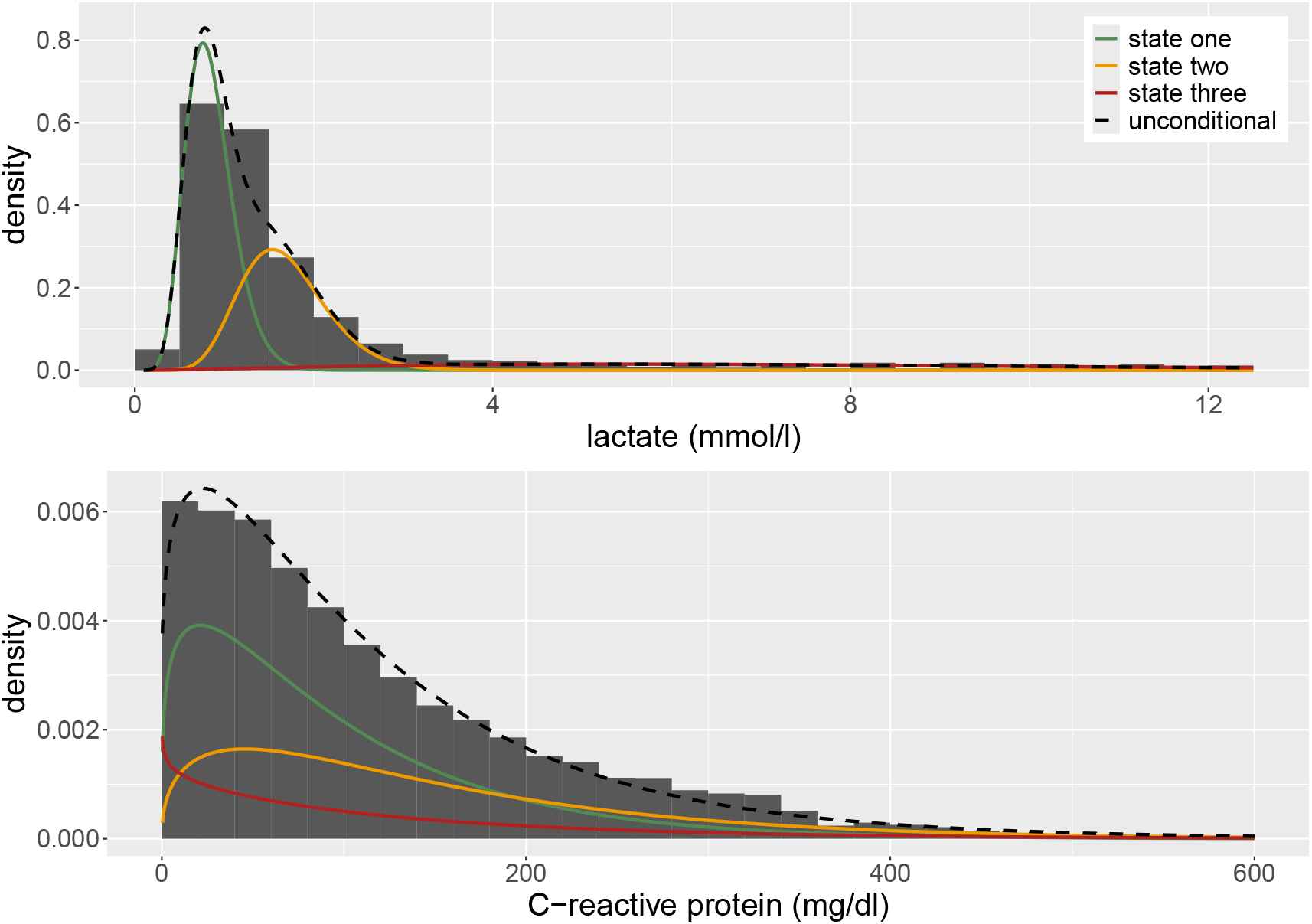
Histograms (shown in dark gray) and estimated state-dependent distributions of lactate (top) and C-reactive protein (bottom). For both laboratory values, we assumed a gamma distribution. The three state-dependent distributions are weighted by the proportion of time spent in the respective state derived from the local state probabilities. The unconditional density is given as black dashed line.

**Table A4:**
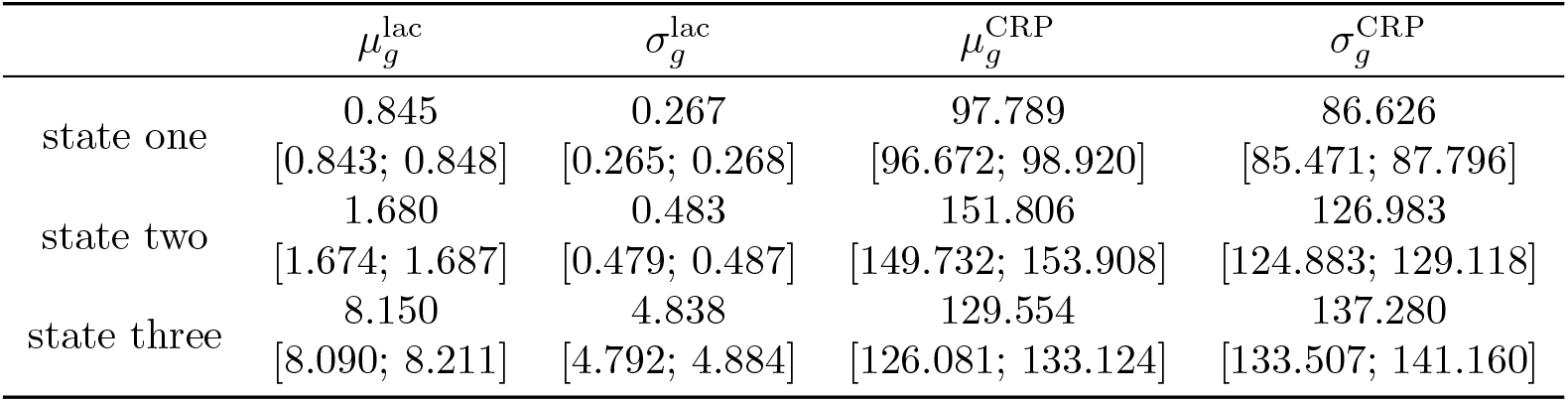
Estimated parameters of the state-dependent processes of lactate and creatinine. Their respective 95 % CIs are given in brackets.

**Table A5:**
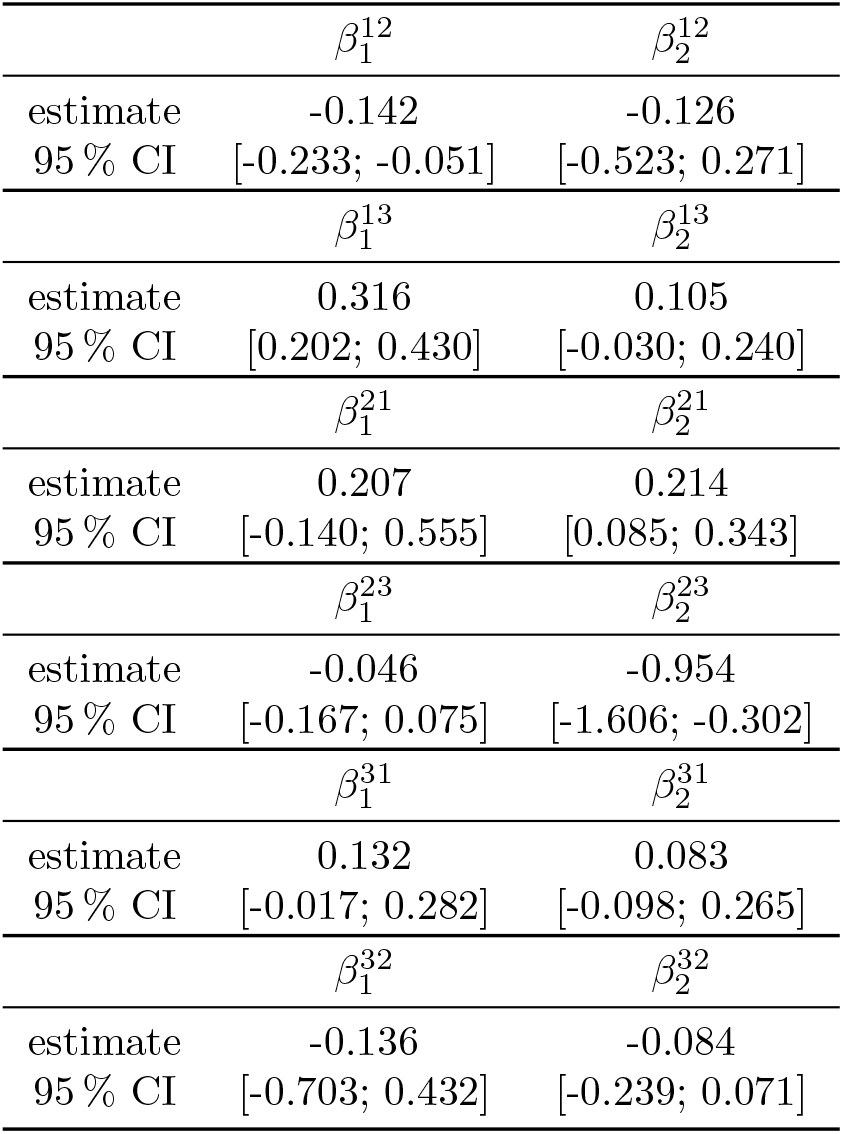
Estimated beta coefficients for the state transition intensities. Their respective 95 % CIs are given in brackets.

| | $\beta_1^{12}$ | $\beta_2^{12}$ |
| --- | --- | --- |
| estimate | -0.142 | -0.126 |
| 95 % CI | [-0.233; -0.051] | [-0.523; 0.271] |
| | $\beta_1^{13}$ | $\beta_2^{13}$ |
| estimate | 0.316 | 0.105 |
| 95 % CI | [0.202; 0.430] | [-0.030; 0.240] |
| | $\beta_1^{21}$ | $\beta_2^{21}$ |
| estimate | 0.207 | 0.214 |
| 95 % CI | [-0.140; 0.555] | [0.085; 0.343] |
| | $\beta_1^{23}$ | $\beta_2^{23}$ |
| estimate | -0.046 | -0.954 |
| 95 % CI | [-0.167; 0.075] | [-1.606; -0.302] |
| | $\beta_1^{31}$ | $\beta_2^{31}$ |
| estimate | 0.132 | 0.083 |
| 95 % CI | [-0.017; 0.282] | [-0.098; 0.265] |
| | $\beta_1^{32}$ | $\beta_2^{32}$ |
| estimate | -0.136 | -0.084 |
| 95 % CI | [-0.703; 0.432] | [-0.239; 0.071] |

### A.4 Estimated Beta Coefficients for the State Transition Intensities

**Table A6:** Estimated coefficients for the groups of the RE for the state transition intensities. Their respective 95 % CIs are given in brackets.

| | $\beta_0^{121}$ | $\beta_0^{122}$ | $\beta_0^{123}$ | $\beta_0^{124}$ |
| --- | --- | --- | --- | --- |
| estimate | -7.583 | -9.928 | -7.368 | -8.330 |
| 95 % CI | [-7.893; -7.274] | [-10.114; -9.741] | [-7.565; -7.171] | [-8.437; -8.224] |
| | $\beta_0^{131}$ | $\beta_0^{132}$ | $\beta_0^{133}$ | $\beta_0^{134}$ |
| estimate | -12.589 | -14.635 | -2.950 | -15.116 |
| 95 % CI | [-13.244; -11.933] | [-18.094; -11.176] | [-3.351; -2.550] | [-23.621; -6.611] |
| | $\beta_0^{211}$ | $\beta_0^{212}$ | $\beta_0^{213}$ | $\beta_0^{214}$ |
| estimate | -10.907 | -8.465 | -9.661 | -9.633 |
| 95 % CI | [-11.270; -10.543] | [-8.642; -8.287] | [-10.503; -8.819] | [-9.784; -9.482] |
| | $\beta_0^{231}$ | $\beta_0^{232}$ | $\beta_0^{233}$ | $\beta_0^{234}$ |
| estimate | -12.533 | -8.335 | -5.069 | -7.035 |
| 95 % CI | [-13.426; -11.641] | [-8.481; -8.188] | [-5.611; -4.528] | [-7.183; -6.887] |
| | $\beta_0^{311}$ | $\beta_0^{312}$ | $\beta_0^{313}$ | $\beta_0^{314}$ |
| estimate | -14.576 | -12.952 | -7.124 | -12.904 |
| 95 % CI | [-19.396; -9.755] | [-15.688; -10.216] | [-7.573; -6.674] | [-13.460; -12.348] |
| | $\beta_0^{321}$ | $\beta_0^{322}$ | $\beta_0^{323}$ | $\beta_0^{324}$ |
| estimate | -9.192 | -9.808 | -11.186 | -10.110 |
| 95 % CI | [-9.763; -8.621] | [-9.943; -9.673] | [-14.599; -7.773] | [-10.313; -9.906] |

### A.5 Estimated Coefficients for the Groups of the Random Effect

**Table A7:** Expected proportion of time spent in the three different states for the four groups of the discrete-valued RE (i.e., RE_1_ to RE_4_) assuming mean-aged female patient.

|  | non-septic state | sepsis-related state | septic shock-related state |
| --- | --- | --- | --- |
| first group | 51.7 % | 2.6 % | 45.7 % |
| second group | 8.4 % | 60.0 % | 31.6 % |
| third group | 81.0 % | 17.9 % | 1.1 % |
| fourth group | 65.9 % | 30.3 % | 3.8 % |

### A.6 Stationary Distribution for the Groups of the Discrete-valued Random Effect

### A.7 Effect of Age and Sex on the Stationary Distribution

**Table A8:** Expected proportion of time spent in the three different states for the youngest and oldest male and female patients in our dataset assuming all of them to belong to the fourth group of the RE.

|  | non-septic state | sepsis-related state | septic shock-related state |
| --- | --- | --- | --- |
| youngest male patient | 85.9 % | 13.2 % | 0.9 % |
| oldest male patient | 65.8 % | 32.6 % | 1.6 % |
| youngest female patient | 80.1 % | 17.2 % | 2.7 % |
| oldest female patient | 55.1 % | 40.5 % | 4.4 % |

### A.8 Effect of Age and Sex on the State Transition Intensities

#### A.8.1 First Group

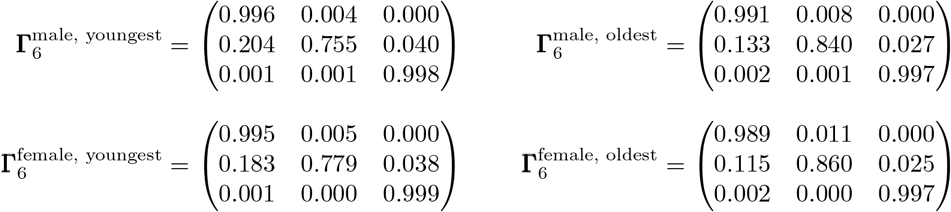

**Table A9:** Dwell times (in hours) in the three different states of the youngest and oldest male and female patients in our dataset assuming all of them to belong to the first group of the RE.

|  | non-septic state | sepsis-related state | septic shock-related state |
| --- | --- | --- | --- |
| youngest male patient | 1268.4 | 21.3 | 2837.9 |
| oldest male patient | 635.0 | 34.3 | 1843.7 |
| youngest female patient | 1027.4 | 24.0 | 4519.6 |
| oldest female patient | 495.7 | 39.6 | 2092.7 |

#### A.8.2 Second Group

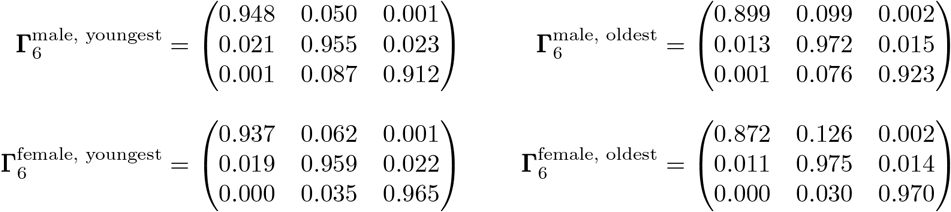

**Table A10:**
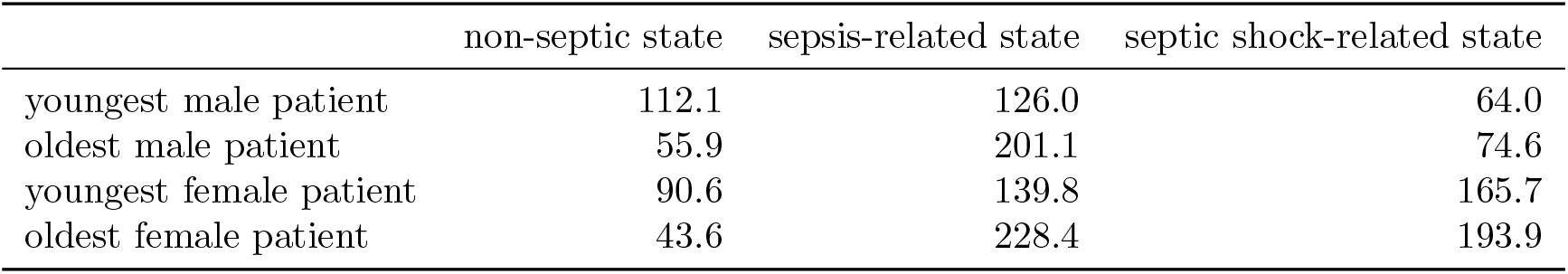
Dwell times (in hours) in the three different states of the youngest and oldest male and female patients in our dataset assuming all of them to belong to the second group of the RE.

|  | non-septic state | sepsis-related state | septic shock-related state |
| --- | --- | --- | --- |
| youngest male patient | 112.1 | 126.0 | 64.0 |
| oldest male patient | 55.9 | 201.1 | 74.6 |
| youngest female patient | 90.6 | 139.8 | 165.7 |
| oldest female patient | 43.6 | 228.4 | 193.9 |

#### A.8.3 Third Group

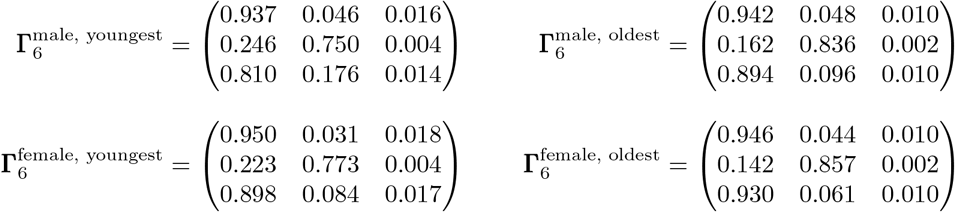

**Table A11:** Dwell times (in hours) in the three different states of the youngest and oldest male and female patients in our dataset assuming all of them to belong to the third group of the RE.

|  | non-septic state | sepsis-related state | septic shock-related state |
| --- | --- | --- | --- |
| youngest male patient | 24.4 | 20.2 | 0.5 |
| oldest male patient | 15.3 | 32.5 | 0.2 |
| youngest female patient | 22.2 | 22.8 | 0.5 |
| oldest female patient | 13.4 | 37.8 | 0.2 |

### A.9 Local State Decoding

**Figure A7:**
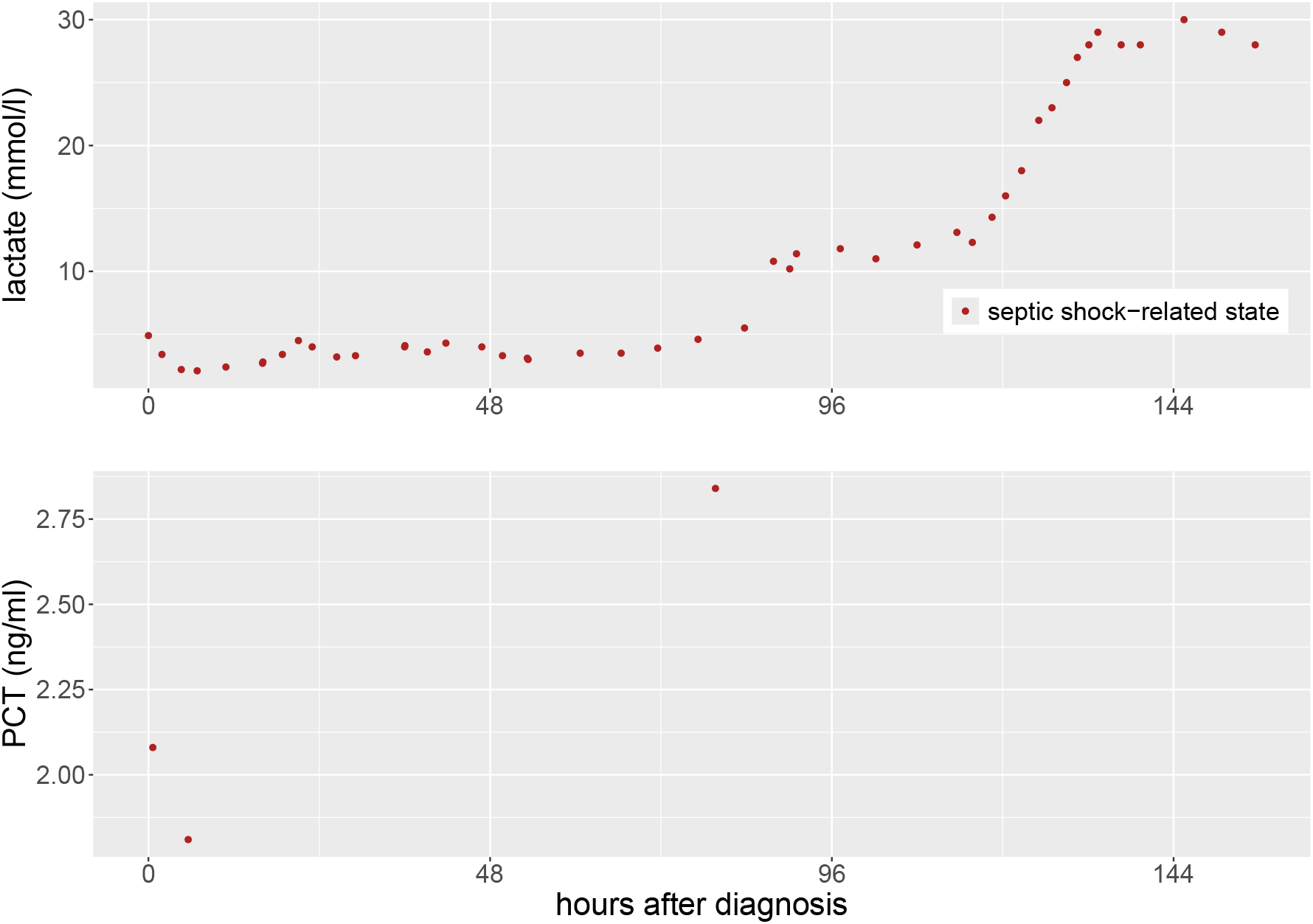
Example of a decoded time series of one stay. This patient never switches between the different states but remains in the septic shock-related state for the entire period. As initial antibiotic treatment, the patient received ‘Ampicillin/Sulbactam-Clarithromycin’. After three days, the therapy was adapted to ‘Piperacillin/Tazobactam’.

#### A.10 Deriving the Effectiveness of Antibiotic Therapies Using LRM

##### A.10.1 Full Dataset

**Figure A8:**
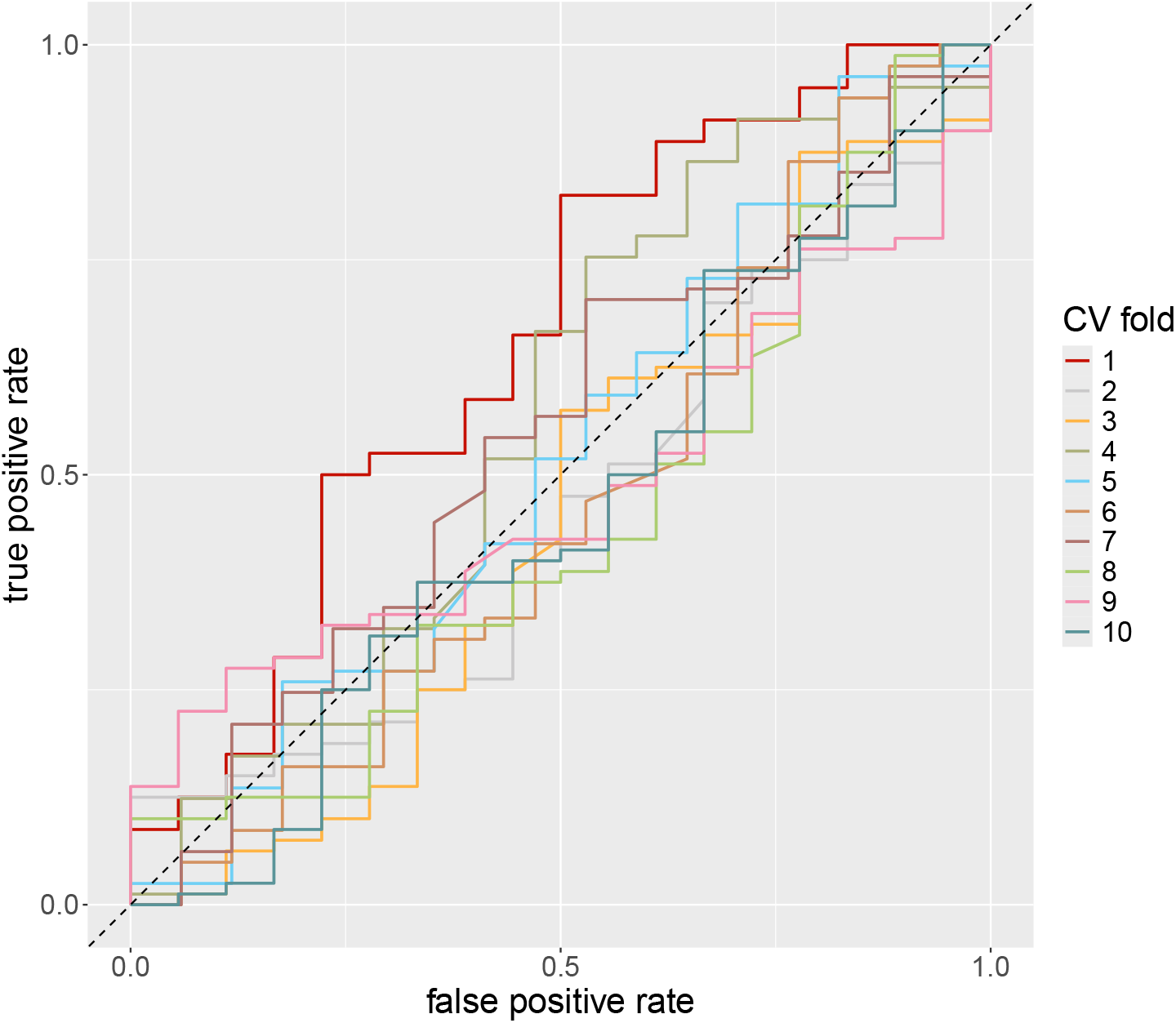
ROC curves of the LRM for the full dataset used to quantify the effectiveness of administered antibiotics across ten folds of CV.

#### A.10.2 Reduced Dataset

**Figure A9:**
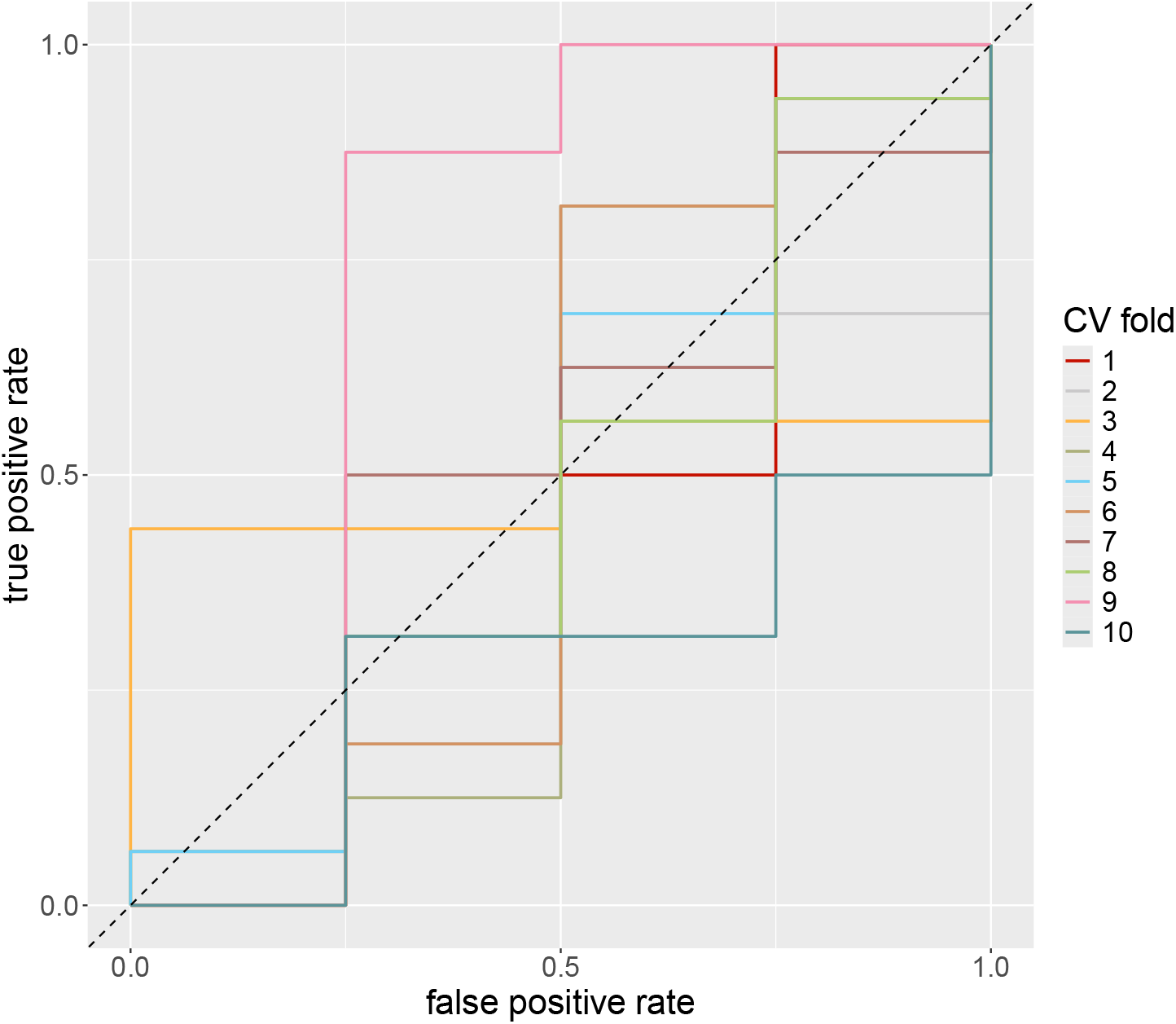
ROC curves of the LRM for the reduced dataset used to quantify the effectiveness of administered antibiotics across ten folds of CV.

## Notes

### Competing Interest Statement

The authors have declared no competing interest.

### Author Declarations

Ethics Committee of the Westphalia-Lippe Medical Association and the University of Muenster gave ethical approval for this work (case number: 2021-699-f-S).

## References

A. M. Alaa, S. Hu, and M. van der Schaar. Learning from Clinical Judgments: Semi-Markov-Modulated Marked Hawkes Processes for Risk Prognosis. In D. Precup and Y. W. Teh, editors, Proceedings of the 34th International Conference on Machine Learning, volume 70 of Proceedings of Machine Learning Research, pages 60–69. PMLR, 2017.

S. E. Allen and J. L. Holm. Lactate: Physiology and Clinical Utility. Journal of Veterinary Emergency and Critical Care, 18(2):123–132, 2008. ISSN 1476-4431. doi: 10.1111/j.1476-4431.2008.00286.x.

R. Amoros, R. King, H. Toyoda, T. Kumada, P. J. Johnson, and T. G. Bird. A Continuous-time Hidden Markov Model for Cancer Surveillance Using Serum Biomarkers With Application to Hepatocellular Carcinoma. METRON, 77:67–86, 2019. ISSN 2281-695X. doi: 10.1007/s40300-019-00151-8.

D. C. Angus and T. van der Poll. Severe Sepsis and Septic Shock. New England Journal of Medicine, 369(9):840–851, 2013. ISSN 1533-4406. doi: 10.1056/nejmra1208623.

A. K. Aslar, M. A. Kuzu, A. H. Elhan, A. Tanik, and S. Hengirmen. Admission Lactate Level and the APACHE II Score are the Most Useful Predictors of Prognosis Following Torso Trauma. Injury, 35(8):746–752, 2004. ISSN 0020-1383. doi: 10.1016/j.injury.2003.09.030.

J. Bakker, M. W. N. Nijsten, and T. C. Jansen. Clinical Use of Lactate Monitoring in Critically Ill Patients. Annals of Intensive Care, 3, 12, 2013. ISSN 2110-5820. doi: 10.1186/2110-5820-3-12.

C. Balci, H. Sungurtekin, E. Gürses, U. Sungurtekin, and B. Kaptanoğlu. Usefulness of Procalcitonin for Diagnosis of Sepsis in the Intensive Care Unit. Critical Care, 7:85–90, 2003. ISSN 1364-8535. doi: 10.1186/cc1843.

K. L. Becker, R. Snider, and E. S. Nylen. Procalcitonin Assay in Systemic Inflammation, Infection, and Sepsis: Clinical Utility and Limitations. Critical Care Medicine, 36(3):941–952, 2008. ISSN 0090-3493. doi: 10.1097/ccm.0b013e318165babb.

A. D. Bendala Estrada, J. Calderón Parra, E. Fernández Carracedo, A. Muiño Míguez, A. Ramos Martínez, E. Muñez Rubio, M. Rubio-Rivas, P. Agudo, F. Arnalich Fernández, V. Estrada Perez, M. L. Taboada Martínez, A. Crestelo Vieitez, P. M. Pesqueira Fontan, M. Bustamante, S. J. Freire, I. Oriol-Bermúdez, A. Artero, J. Olalla Sierra, M. Areses Manrique, H. F. J. Carrasco-Sánchez, V. C. Vento, G. M. García García, P. Cubero-Morais, J.-M. Casas-Rojo, and J. M. Núñez-Cortés. Inadequate Use of Antibiotics in the Covid-19 Era: Effectiveness of Antibiotic Therapy. BMC Infectious Diseases, 21, 1144, 2021. ISSN 1471-2334. doi: 10.1186/s12879-021-06821-1.

P. Blin, S. Blazejewski, S. Lignot, R. Lassalle, M. Bernard, D. Jayles, H. Théophile, J. Bénichou, J. Demeaux, D. Ebbo, J. Franck, Y. Moride, D. Peyramond, B. Rouveix, M. Sturkenboom, P. Gehanno, C. Droz, and N. Moore. Effectiveness of Antibiotics for Acute Sinusitis in Real-life Medical Practice. British Journal of Clinical Pharmacology, 70(3):418–428, 2010. ISSN 1365-2125. doi: 10.1111/j.1365-2125.2010.03710.x.

A. Bureau, S. Shiboski, and J. P. Hughes. Applications of Continuous Time Hidden Markov Models to the Study of Misclassified Disease Outcomes. Statistics in Medicine, 22(3):441–462, 2003. ISSN 1097-0258. doi: 10.1002/sim.1270.

A. R. Burrell, M. McLaws, M. Fullick, R. B. Sullivan, and D. Sindhusake. SEPSIS KILLS: Early Intervention Saves Lives. Medical Journal of Australia, 204(2):73–73, 2016. ISSN 1326-5377. doi: 10.5694/mja15.00657.

O. Cerović, V. Golubović, A. Špec Marn, B. Kremžar, and G. Vidmar. Relationship Between Injury Severity and Lactate Levels in Severely Injured Patients. Intensive Care Medicine, 29:1300–1305, 2003. ISSN 1432-1238. doi: 10.1007/s00134-003-1753-8.

P. E. Charles, C. Tinel, S. Barbar, S. Aho, S. Prin, J. M. Doise, N. O. Olsson, B. Blettery, and J. P. Quenot. Procalcitonin Kinetics Within the First Days of Sepsis: Relationship With the Appropriateness of Antibiotic Therapy and the Outcome. Critical Care, 13(2, R38), 2009. ISSN 1364-8535. doi: 10.1186/cc7751.

S. L. DeRuiter, R. Langrock, T. Skirbutas, J. A. Goldbogen, J. Calambokidis, A. S. Friedlaender, and B. L. Southall. A Multivariate Mixed Hidden Markov Model for Blue Whale Behaviour and Responses to Sound Exposure. The Annals of Applied Statistics, 11(1):362–392, 2017. ISSN 1932-6157. doi: 10.1214/16-aoas1008.

Deutsche Sepsis-Gesellschaft, Deutsche Gesellschaft für Anästhesiologie und Intensivmedizin, and Deutsche Interdisziplinäre Vereinigung für Intensiv-und Notfallmedizin et al. [AWMF-Register-Nr.: 079-001] S3-Leitlinie: Sepsis – Prävention, Diagnose, Therapie und Nachsorge – Update 2025. Langfassung. Version: 4.0, Stand: 30.04.2025, 2025. URL https://register.awmf.org/de/leitlinien/detail/079-001.

L. Evans, A. Rhodes, W. Alhazzani, M. Antonelli, C. M. Coopersmith, C. French, F. R. Machado, L. Mcintyre, M. Ostermann, H. C. Prescott, C. Schorr, S. Simpson, W. J. Wiersinga, F. Alshamsi, D. C. Angus, Y. Arabi, L. Azevedo, R. Beale, G. Beilman, E. Belley-Cote, L. Burry, M. Cecconi, J. Centofanti, A. Coz Yataco, J. de Waele, R. P. Dellinger, K. Doi, B. Du, E. Estenssoro, R. Ferrer, C. Gomersall, C. Hodgson, M. H. Møller, T. Iwashyna, S. Jacob, R. Kleinpell, M. Klompas, Y. Koh, A. Kumar, A. Kwizera, S. Lobo, H. Masur, S. McGloughlin, S. Mehta, Y. Mehta, M. Mer, M. Nunnally, S. Oczkowski, T. Osborn, E. Papathanassoglou, A. Perner, M. Puskarich, J. Roberts, W. Schweickert, M. Seckel, J. Sevransky, C. L. Sprung, T. Welte, J. Zimmerman, and M. Levy. Surviving Sepsis Campaign: International Guidelines for Management of Sepsis and Septic Shock 2021. Intensive care medicine, 47(11):1181–1247, 2021. ISSN 1432-1238. doi: 10.1007/s00134-021-06506-y.

G. Feretzakis, A. Sakagianni, E. Loupelis, D. Kalles, N. Skarmoutsou, M. Martsoukou, C. Christopoulos, M. Lada, S. Petropoulou, A. Velentza, S. Michelidou, R. Chatzikyriakou, and E. Dimitrellos. Machine Learning for Antibiotic Resistance Prediction: A Prototype Using Off-the-Shelf Techniques and Entry-Level Data to Guide Empiric Antimicrobial Therapy. Healthcare Informatics Research, 27(3):214–221, 2021. ISSN 2093-369X. doi: 10.4258/hir.2021.27.3.214.

R. R. Filho, L. L. Rocha, T. D. Corrêa, C. M. S. Pessoa, G. Colombo, and M. S. C. Assuncao. Blood Lactate Levels Cutoff and Mortality Prediction in Sepsis—Time for a Reappraisal? a Retrospective Cohort Study. Shock, 46(5):480–485, 2016. ISSN 1073-2322. doi: 10.1097/shk.0000000000000667.

J.-O. Fischer. LaMa: Fast Numerical Maximum Likelihood Estimation for Latent Markov Models, 2025. URL https://CRAN.R-project.org/package=LaMa. R package version 2.0.6.

C. Fleischmann, D. O. Thomas–Rueddel, M. Hartmann, C. S. Hartog, T. Welte, S. Heublein, U. Dennler, and K. Reinhart. Hospital Incidence and Mortality Rates of Sepsis: An Analysis of Hospital Episode (DRG) Statistics in Germany From 2007 to 2013. Deutsches Ärzteblatt international, 113:159–166, 2016. ISSN 1866-0452. doi: 10.3238/arztebl.2016.0159.

G. D. Forney. The Viterbi Algorithm. Proceedings of the IEEE, 61(3):268–278, 1973. ISSN 0018-9219. doi: 10.1109/proc.1973.9030.

R. Glennie, T. Adam, V. Leos-Barajas, T. Michelot, T. Photopoulou, and B. T. McClintock. Hidden Markov models: Pitfalls and Opportunities in Ecology. Methods in Ecology and Evolution, 14 (1):43–56, 2023. ISSN 2041-210X. doi: 10.1111/2041-210x.13801.

E. Gultepe, J. P. Green, H. Nguyen, J. Adams, T. Albertson, and I. Tagkopoulos. From Vital Signs to Clinical Outcomes for Patients With Sepsis: a Machine Learning Basis for a Clinical Decision Support System. Journal of the American Medical Informatics Association, 21(2):315–325, 2014. ISSN 1527-974X. doi: 10.1136/amiajnl-2013-001815.

S. Hagel and F. Brunkhorst. Sepsis. Intensivmedizin und Notfallmedizin, 48(1):57–73, 2011. ISSN 1435-1420. doi: 10.1007/s00390-010-0249-3.

C. H. Jackson, L. D. Sharples, S. G. Thompson, S. W. Duffy, and E. Couto. Multistate Markov Models for Disease Progression With Classification Error. Journal of the Royal Statistical Society Series D: The Statistician, 52(2):193–209, 2003. ISSN 1467-9884. doi: 10.1111/1467-9884.00351.

K. Kristensen. RTMB: ‘R’ Bindings for ‘TMB’, 2025. URL https://CRAN.R-project.org/package=RTMB. R package version 1.8.

J. A. Kruse, S. A. J. Zaidi, and R. W. Carlson. Significance of Blood Lactate Levels in Critically Ill Patients with Liver Disease. The American Journal of Medicine, 83(1):77–82, 1987. ISSN 0002-9343. doi: 10.1016/0002-9343(87)90500-6.

J. M. Lange, R. A. Hubbard, L. Y. T. Inoue, and V. N. Minin. A Joint Model for Multistate Disease Processes and Random Informative Observation Times, with Applications to Electronic Medical Records Data. Biometrics, 71(1):90–101, 2015. ISSN 1541-0420. doi: 10.1111/biom.12252.

O. Lewin-Epstein, S. Baruch, L. Hadany, G. Y. Stein, and U. Obolski. Predicting Antibiotic Resistance in Hospitalized Patients by Applying Machine Learning to Electronic Medical Records. Clinical Infectious Diseases, 72(11):e848–e855, 2021. ISSN 1537-6591. doi: 10.1093/cid/ciaa1576.

Y.-Y. Liu, S. Li, F. Li, L. Song, and J. M. Rehg. Efficient Learning of Continuous-Time Hidden Markov Models for Disease Progression. In C. Cortes, N. Lawrence, D. Lee, M. Sugiyama, and aR. Garnett, editors, Advances in Neural Information Processing Systems, volume 28. Curran Associates, Inc., 2015.

J. F. S. Logman, J. Stephens, B. Heeg, S. Haider, J. Cappelleri, D. Nathwani, A. Tice, and B. A. van Hout. Comparative Effectiveness of Antibiotics for the Treatment of MRSA Complicated Skin and Soft Tissue Infections. Current Medical Research and Opinion, 26(7):1565–1578, 2010. ISSN 1473-4877. doi: 10.1185/03007995.2010.481251.

A. Luzzani, E. Polati, R. Dorizzi, A. Rungatscher, R. Pavan, and A. Merlini. Comparison of Procalcitonin and C-reactive Protein as Markers of Sepsis. Critical Care Medicine, 31(6):1737–1741, 2003. ISSN 0090-3493. doi: 10.1097/01.ccm.0000063440.19188.ed.

H. Marchi, S. Schmiegel, R. Borgstedt, S. Rehberg, and C. Fuchs. Hybrid Health Recommender Systems for Antibiotic Prescription in Sepsis Patients: Statistical Method Development and Evaluation. medRxiv, 2025. doi: 10.1101/2025.11.21.25340749.

J. L. Martínez, F. Baquero, and D. I. Andersson. Predicting Antibiotic Resistance. Nature Reviews Microbiology, 5:958–965, 2007. ISSN 1740-1534. doi: 10.1038/nrmicro1796.

A. Maruotti and T. Rydén. A Semiparametric Approach to Hidden Markov Models Under Longitudinal Observations. Statistics and Computing, 19, 381, 2009. ISSN 1573-1375. doi: 10.1007/s11222-008-9099-2.

F. B. Mayr, S. Yende, and D. C. Angus. Epidemiology of Severe Sepsis. Virulence, 5(1):4–11, 2014. ISSN 2150-5608. doi: 10.4161/viru.27372.

A. E. McKellar, R. Langrock, J. R. Walters, and D. C. Kesler. Using Mixed Hidden Markov Models to Examine Behavioral States in a Cooperatively Breeding Bird. Behavioral Ecology, 26 (1):148–157, 2015. ISSN 1045-2249. doi: 10.1093/beheco/aru171.

M. Meisner, K. Tschaikowsky, T. Palmaers, and J. Schmidt. Comparison of Procalcitonin (PCT) and C-reactive Protein (CRP) Plasma Concentrations at Different SOFA Scores During the Course of Sepsis and MODS. Critical Care, 3, 45, 1999. ISSN 1364-8535. doi: 10.1186/cc306.

S. Mews, B. Surmann, L. Hasemann, and S. Elkenkamp. Markov-modulated Marked Poisson Processes for Modeling Disease Dynamics Based on Medical Claims Data. Statistics in Medicine, 42 (21):3804–3815, 2023. ISSN 1097-0258. doi: 10.1002/sim.9832.

M. E. Mikkelsen, A. N. Miltiades, D. F. Gaieski, M. Goyal, B. D. Fuchs, C. V. Shah, S. L. Bellamy, and J. D. Christie. Serum Lactate is Associated With Mortality in Severe Sepsis Independent of Organ Failure and Shock*. Critical Care Medicine, 37(5):1670–1677, 2009. ISSN 0090-3493. doi: 10.1097/ccm.0b013e31819fcf68.

C. J. L. Murray, K. S. Ikuta, F. Sharara, L. Swetschinski, G. Robles Aguilar, A. Gray, C. Han, C. Bisignano, P. Rao, E. Wool, S. C. Johnson, A. J. Browne, M. G. Chipeta, F. Fell, S. Hackett, G. Haines-Woodhouse, B. H. Kashef Hamadani, E. A. P. Kumaran, B. McManigal, S. Achalapong, R. Agarwal, S. Akech, S. Albertson, J. Amuasi, J. Andrews, A. Aravkin, E. Ashley, F.-X. Babin, F. Bailey, S. Baker, B. Basnyat, A. Bekker, R. Bender, J. A. Berkley, A. Bethou, J. Bielicki, S. Boonkasidecha, J. Bukosia, C. Carvalheiro, C. Castañeda-Orjuela, V. Chansamouth, S. Chaurasia, S. Chiurchiú, F. Chowdhury, R. Clotaire Donatien, A. J. Cook, B. Cooper, T. R. Cressey, E. Criollo-Mora, M. Cunningham, S. Darboe, N. P. J. Day, M. De Luca, K. Dokova, A. Dramowski, S. J. Dunachie, T. Duong Bich, T. Eckmanns, D. Eibach, A. Emami, N. Feasey, N. Fisher-Pearson, K. Forrest, C. Garcia, D. Garrett, P. Gastmeier, A. Z. Giref, R. C. Greer, V. Gupta, S. Haller, A. Haselbeck, S. I. Hay, M. Holm, S. Hopkins, Y. Hsia, K. C. Iregbu, J. Jacobs, D. Jarovsky, F. Javanmardi, A. W. J. Jenney, M. Khorana, S. Khusuwan, N. Kissoon, E. Kobeissi, T. Kostyanev, F. Krapp, R. Krumkamp, A. Kumar, H. H. Kyu, C. Lim, K. Lim, D. Limmathurotsakul, M. J. Loftus, M. Lunn, J. Ma, A. Manoharan, F. Marks, J. May, M. Mayxay, N. Mturi, T. Munera-Huertas, P. Musicha, L. A. Musila, M. M. Mussi-Pinhata, R. N. Naidu, T. Nakamura, R. Nanavati, S. Nangia, P. Newton, C. Ngoun, A. Novotney, D. Nwakanma, C. W. Obiero, T. J. Ochoa, A. Olivas-Martinez, P. Olliaro, E. Ooko, E. Ortiz-Brizuela, P. Ounchanum, G. D. Pak, J. L. Paredes, A. Y. Peleg, C. Perrone, T. Phe, K. Phommasone, N. Plakkal, A. Ponce-de Leon, M. Raad, T. Ramdin, S. Rattanavong, A. Riddell, T. Roberts, J. V. Robotham, A. Roca, V. D. Rosenthal, K. E. Rudd, N. Russell, H. S. Sader, W. Saengchan, J. Schnall, J. A. G. Scott, S. Seekaew, M. Sharland, M. Shivamallappa, J. Sifuentes-Osornio, A. J. Simpson, N. Steenkeste, A. J. Stewardson, T. Stoeva, N. Tasak, A. Thaiprakong, G. Thwaites, C. Tigoi, C. Turner, P. Turner, H. R. van Doorn, S. Velaphi, A. Vongpradith, M. Vongsouvath, H. Vu, T. Walsh, J. L. Walson, S. Waner, T. Wangrangsimakul, P. Wannapinij, T. Wozniak, T. E. M. W. Young Sharma, K. C. Yu, P. Zheng, B. Sartorius, A. D. Lopez, A. Stergachis, C. Moore, C. Dolecek, and M. Naghavi. Global Burden of Bacterial Antimicrobial Resistance in 2019: a Systematic Analysis. The Lancet, 399(10325):629–655, 2022. ISSN 0140-6736. doi: 10.1016/s0140-6736(21)02724-0.

J. D. Parente, K. Möller, G. M. Shaw, and J. G. Chase. Hidden Markov Models for Sepsis Classification. IFAC-PapersOnLine, 51(27):110–115, 2018. ISSN 2405-8963. doi: 10.1016/j.ifacol.2018.11.658.

T. A. Patterson, A. Parton, R. Langrock, P. G. Blackwell, L. Thomas, and R. King. Statistical Modelling of Individual Animal Movement: an Overview of Key Methods and a Discussion of Practical Challenges. AStA Advances in Statistical Analysis, 101:399–438, 2017. ISSN 1863-818X. doi: 10.1007/s10182-017-0302-7.

L. Pruinelli, B. L. Westra, P. Yadav, A. Hoff, M. Steinbach, V. Kumar, C. W. Delaney, and G. Simon. Delay Within the 3-Hour Surviving Sepsis Campaign Guideline on Mortality for Patients With Severe Sepsis and Septic Shock*. Critical Care Medicine, 46(4):500–505, 2018. ISSN 0090-3493. doi: 10.1097/ccm.0000000000002949.

M. A. Puskarich, J. A. Kline, R. L. Summers, and A. E. Jones. Prognostic Value of Incremental Lactate Elevations in Emergency Department Patients With Suspected Infection. Academic Emergency Medicine, 19(8):983–985, 2012. ISSN 1553-2712. doi: 10.1111/j.1553-2712.2012.01404.x.

R Core Team. R: A Language and Environment for Statistical Computing. R Foundation for Statistical Computing, Vienna, Austria, 2024. URL https://www.R-project.org/.

M. S. Rangel-Frausto, D. Pittet, T. Hwang, R. F. Woolson, and R. P. Wenzel. The Dynamics of Disease Progression in Sepsis: Markov Modeling Describing the Natural History and the Likely Impact of Effective Antisepsis Agents. Clinical Infectious Diseases, 27(1):185–190, 1998. ISSN 1537-6591. doi: 10.1086/514630.

P.-M. Roger, C. Labate, S. Serre, C. Zumbo, L. Valério, H. Bonnet, A. Jurado, N. Bélé, and P. Brofferio. Factors Associated With Effective Reassessment of Antibiotic Therapy on Day 3. Médecine et Maladies Infectieuses, 43(3):123–127, 2013. ISSN 0399-077X. doi: 10.1016/j.medmal.2012.12.007.

S. M. Ryoo and W. Y. Kim. Clinical Applications of Lactate Testing in Patients With Sepsis and Septic Shock. Journal of Emergency and Critical Care Medicine, 2(2), 2018. ISSN 2521-3563. doi: 10.21037/jeccm.2018.01.13.

I. Samsudin and S. D. Vasikaran. Clinical Utility and Measurement of Procalcitonin. The Clinical Biochemist Reviews, 38(2):59–68, 2017.

S. Schmiegel, H. Marchi, P. Hege, S. Elkenkamp, J. Duevel, C. Düsing, W. Greiner, S. S. Scholz, D. Witzke, J. J. Tebbe, M. Wehmeier, O. Kaup, R. Borgstedt, S. Rehberg, P. Cimiano, and C. Fuchs. Development Process of a Clinical Decision Support System for Empiric Antibiotic Therapies in Patients With Sepsis: Case Study. JMIR Medical Informatics, 14:e79929–e79929, 2026. ISSN 2291-9694. doi: 10.2196/79929.

C. W. Seymour, F. Gesten, H. C. Prescott, M. E. Friedrich, T. J. Iwashyna, G. S. Phillips, S. Lemeshow, T. Osborn, K. M. Terry, and M. M. Levy. Time to Treatment and Mortality during Mandated Emergency Care for Sepsis. New England Journal of Medicine, 376(23):2235–2244, 2017. ISSN 1533-4406. doi: 10.1056/nejmoa1703058.

H. Shamsudin, U. K. Yusof, A. Jayalakshmi, and M. N. Akmal Khalid. Combining Oversampling and Undersampling Techniques for Imbalanced Classification: A Comparative Study Using Credit Card Fraudulent Transaction Dataset. In 2020 IEEE 16th International Conference on Control & Automation (ICCA), pages 803–808. IEEE, 2020. doi: 10.1109/icca51439.2020.9264517.

M. Singer, C. S. Deutschman, C. W. Seymour, M. Shankar-Hari, D. Annane, M. Bauer, R. Bellomo, G. R. Bernard, J.-D. Chiche, C. M. Coopersmith, R. S. Hotchkiss, M. M. Levy, J. C. Marshall, G. S. Martin, S. M. Opal, G. D. Rubenfeld, T. van der Poll, J.-L. Vincent, and D. C. Angus. The Third International Consensus Definitions for Sepsis and Septic Shock (Sepsis-3). JAMA, 315(8):801–810, 2016. doi: 10.1001/jama.2016.0287.

A. V. Towner, V. Leos-Barajas, R. Langrock, R. S. Schick, M. J. Smale, T. Kaschke, O. J. D. Jewell, and Y. P. Papastamatiou. Sex-specific and Individual Preferences for Hunting Strategies in White Sharks. Functional Ecology, 30(8):1397–1407, 2016. ISSN 1365-2435. doi: 10.1111/1365-2435.12613.

S. Trzeciak, R. P. Dellinger, M. E. Chansky, R. C. Arnold, C. Schorr, B. Milcarek, S. M. Hollenberg, and J. E. Parrillo. Serum Lactate as a Predictor of Mortality in Patients With Infection. Intensive Care Medicine, 33(6):970–977, 2007. ISSN 1432-1238. doi: 10.1007/s00134-007-0563-9.

A. Viterbi. Error Bounds for Convolutional Codes and an Asymptotically Optimum Decoding Algorithm. IEEE Transactions on Information Theory, 13(2):260–269, 1967. ISSN 1557-9654. doi: 10.1109/tit.1967.1054010.

K. T. Whang, P. M. Steinwald, J. C. White, E. S. Nylen, R. H. Snider, G. L. Simon, R. L. Goldberg, and K. L. Becker. Serum Calcitonin Precursors in Sepsis and Systemic Inflammation*. The Journal of Clinical Endocrinology & Metabolism, 83(9):3296–3301, 1998. ISSN 1945-7197. doi: 10.1210/jcem.83.9.5129.

Z. Zhang and X. Xu. Lactate Clearance Is a Useful Biomarker for the Prediction of All-Cause Mortality in Critically Ill Patients: A Systematic Review and Meta-Analysis*. Critical Care Medicine, 42(9):2118–2125, 2014. ISSN 0090-3493. doi: 10.1097/ccm.0000000000000405.

W. Zucchini, I. L. MacDonald, and R. Langrock. Hidden Markov Models for Time Series: An Introduction Using R. Chapman and Hall/CRC, 2 edition, 2016. ISBN 9781315372488. doi: 10.1201/b20790.

